# Monitoring of Open Science practices: a survey of 10 major medical journals

**DOI:** 10.1101/2025.11.26.25341061

**Authors:** Constant Vinatier, Ayu Putu Madri Dewi, Guillaume Freyermuth, Margaux Millour, Maud Scheidecker, François-Joseph Arnault, Mathieu Acher, Vladislav Nachev, Tracey L. Weissgerber, Nicholas J. DeVito, Gwénaël Dumont, Gowri Gopalakrishna, Gauthier Le Bartz Lyan, Inge Stegeman, Mariska M. G. Leeflang, Florian Naudet

**Affiliations:** Univ Rennes, CHU Rennes, Inserm, EHESP, Irset (Institut de recherche en santé, environnement et travail) – UMR_S 1085, F-35000 Rennes, France; Department of Epidemiology and Data Science, Amsterdam University Medical Centers, Amsterdam, the Netherlands; Univ Rennes, IRISA, Inria, CNRS, F-35000 Rennes, France; Institut Universitaire de France (IUF), Paris, France; Berlin Institute of Health at Charité – Universitätsmedizin Berlin, QUEST Center for Responsible Research, Berlin, Germany; CNC-UC, Center for Neuroscience and Cell Biology, University of Coimbra, Coimbra, Portugal; CIBB, Center for Innovative Biomedicine and Biotechnology, University of Coimbra, Coimbra, Portugal; Bennett Institute for Applied Data Science, Nuffield Department of Primary Care Health Sciences, University of Oxford, Oxford, United Kingdom; Department of Epidemiology, Faculty of Health, Medicine, and Life Sciences Maastricht University; Department of Otorhinolaryngology and Head & Neck Surgery University Medical Center Utrecht Utrecht The Netherlands.; Brain Center, University Medical Center Utrecht, Utrecht, The Netherlands

## Abstract

**Objective:** To evaluate open science policies of leading general medical journals and the extent to which open science practices are implemented and detectable using automated tools.

**Design:** Cross-sectional audit of journal policies and retrospective observational study of journal articles, with diagnostic accuracy validation of automated screening tools against manual data extraction.

Setting: Annals of Internal Medicine, BMC Medicine, The BMJ, CMAJ, JAMA, JAMA Network Open, The Lancet, Nature Medicine, New England Journal of Medicine, and PLOS Medicine.

Participants:Research articles published between 2020 and 2023 and randomised controlled trials (RCTs) published before and after policy changes regarding registration and data-sharing.

Main outcome measures: Journal policies were assessed using the TOP2025 framework, which rates journals’ transparency and openness standards using the TOP Factor (maximum score 27). At the article level, thirteen core open science practices were examined, including trial registration, protocol sharing, and intention to share data. Nine validated automated tools were applied to detect these practices and compared with manual extraction of 312 articles performed in duplicate. Changes in transparency practices following policy updates were analyzed.

**Results:** Transparency policies varied considerably. TOP Factor scores ranged from 1 (NEJM) to 13 (PLOS Medicine), with many journals’ policies applying primarily to clinical trials rather than all research articles. Only one journal (BMC Medicine) proposed registered reports. At the article level, adoption of open science practices was highest in RCTs: trial registration (99% [95%–100%] vs 68% [55%–79%] in meta-analyses vs 15% [8%–27%] in other research), protocol sharing (95% [90%–98%] vs 67% [54%–78%] vs 19% [11%–32%]), and intention to share data (78% [67%–87%] vs 64% [49%–76%] vs 69% [55%–80%]). Performance across tools was very variable (F1 scores 0.16–1.00). Among all 15,624 research articles, tools detected registration in 20%, protocol sharing in 42%, and intention to share data in 48%, but these were generally underestimated. Policy changes such as mandatory registration or data-sharing requirements were associated with measurable improvements in the corresponding open science practices.

**Conclusions:** Leading medical journals show only partial alignment with TOP2025, with stronger transparency observed for RCTs than for other study types. Triangulation with manual and automated assessments in articles confirms that key open science practices remain suboptimal especially for non-RCTs, highlighting the need for more robust journal policies. Policy changes are associated with observable improvements.

Registration: OSF: https://doi.org/10.17605/OSF.IO/F2VW9

## Introduction

Medical journals play a central role in disseminating biomedical research, which is essential for advancing healthcare and informing clinical decision making. Open science practices, such as study registration, protocol availability, and data sharing, are increasingly recognised as fundamental components of high-quality research (1).

In 2005, the International Committee of Medical Journal Editors (ICMJE) required prospective registration of clinical trials as a condition for publication(2). Progress in other areas, including data sharing, has been slower. The Annals of Internal Medicine first encouraged data sharing in 2007 (3). In 2018, the ICMJE introduced data sharing statements for clinical trials, requiring authors to declare whether and how they would share data (4). Compliance, however, remains limited (5,6).

Some leading journals have adopted stronger measures. The BMJ began encouraging data sharing for clinical trials in 2009 (7), made it mandatory for drug and device trials in 2013 (8), extended the policy to all clinical trials in 2015 (9), and from May 2024 requires data deposition in a public repository (10). PLOS Medicine adopted a similar requirement in 2014 (11). These initiatives, however, remain largely confined to clinical trials. Extending open science practices to other study designs, such as observational research (12), could further strengthen reproducibility. Biomedical journals have also been slow to adopt innovative formats such as registered reports (13), a publication model in which study methods are peer-reviewed and accepted in principle before data collection begins, thereby reducing publication bias. In contrast, registered reports are more common in psychology (14).

The emphasis on transparency aligns with broader reforms in research assessment. The San Francisco Declaration on Research Assessment (DORA) discourages reliance on journal impact factors (15), while the Hong Kong Principles advocate rewarding open and reproducible practices (16). International bodies, including the G7 (17) and UNESCO (18), have made open science a strategic priority and published Principles of Open Science Monitoring (19). The updated Transparency and Openness Promotion (TOP) Guidelines (TOP2025) (20) provide frameworks for evaluating policies. At the article level, the ScreenIT Working Group has developed automated tools to enable large-scale monitoring of open science practices (21). In biomedicine, there is also consensus on which practices should be assessed (22).

Monitoring how journals implement and promote open science is therefore essential to determine how well stated commitments to transparency translate into practice. Systematic monitoring can identify gaps, track progress, and strengthen the credibility and societal value of biomedical research. It may also help counter the influence of predatory journals (23) by highlighting those with suboptimal standards (24). In this study, we aimed to develop a proof-of-concept observatory of open science practices in leading medical journals. Specifically, we sought to evaluate the feasibility of using automated tools to monitor transparency indicators at the article level; to describe current open science policies in those journals; and analyze retrospectively changes in transparency practices over time, particularly following major policy shifts.

## Methods

### Overall approach

By “observatory of open science practices,” we mean a systematic platform that monitors and evaluates the adoption of transparency and reproducibility practices across journals, at the policy level, and their published research articles. As a proof-of-concept assessing feasibility, this report presents a static, retrospective analysis of open science practices in leading medical journals. We audited the journals’ policies using the 2025 TOP Guidelines, evaluated the diagnostic accuracy of several automated tools for detecting transparency indicators at the article level, investigated several technical aspects, including differences in tool performance between XML and PDF file formats, assessed open science practices in recent journal articles using the best-performing tools and evaluated selected practices (intention to share data and registration) before and after the journals’ policy change.

### Journal Selection

We analyzed ten prominent medical journals, purposefully chosen for their international reputation as leading general medical publications. Specialist journals were not included. Consistent with DORA (15), we did not use impact factor for selection. Most included journals were either current or former ICMJE members, such as Annals of Internal Medicine, The BMJ, CMAJ, JAMA, The Lancet, Nature Medicine, New England Journal of Medicine, and PLOS Medicine. Furthermore, two major open access medical journals, JAMA Network Open and BMC Medicine, were also included.

### TOP2025 journal policy audit

Two researchers (CV and APMD) independently extracted information related to the TOP factor criteria for each included journal (20). **Web Appendix 1** lists all items evaluated by TOP 2025. Data extraction relied on either a TOP assessment conducted by the Center for Open Science within the previous year, if available, or on a direct analysis of editorial policies published on each journal’s website. If there was disagreement, a third researcher (FN) made the final decision. Besides TOP2025, we also checked whether the journal or publisher is a DORA signatory. All editorial teams were invited to review our extraction and provide any necessary corrections. We requested feedback within one month, sending reminder communications during the first and third weeks.

### Research articles search strategy

After assessing journal policies, we examined articles published in those journals. Our search strategy aimed to include all types of research articles classified as randomized controlled trials (RCTs), meta-analyses, or other types of research articles, and to exclude congress communications, opinion papers, editorials, and other non-research articles. Publication dates for the articles in the various validation databases and in the observatory are provided in the corresponding sections.

All eligible articles from the selected journals were identified through three distinct PubMed queries, tailored by study type. The search strategies were reviewed by an information specialist and adhered to the peer review of electronic literature search strategies (PRESS) guidelines (25). Detailed search strategies are provided in **Web Appendix 2**. We primarily used PDF files and supplements (in pdf format) for data collection. For The Lancet, where PDFs and supplements were unavailable, we utilized the Elsevier API to download all available XML files containing the print content of the article. **Web Appendix 3** provides detailed information for each journal.

### Diagnostic accuracy of tools to monitor open science practices

#### Open science practices

**Web Appendix 4** details the 19 core open science practices (with 12 « traditional open science practices » and 7 « broader open science practices ») identified for monitoring in biomedicine through prior consensus (22). Of these, we retained 13 practices based on whether the information could be obtained from a published article. Monitored practices include registration, intention to share data, open access publishing, code sharing, protocol sharing, statistical analysis plan sharing, the use of a reporting guideline, presence of a preprint, presence of author contribution, presence of a conflict of interest (COI) statement, reporting the ORCID identifiers of authors and presence of a funding statement. Additionally, for RCT, we monitored the publication 1 year after the end of the study. The reasons for excluding other items from the consensus list of open science practices to be monitored are provided in **Table 1**.

**Table 1:**
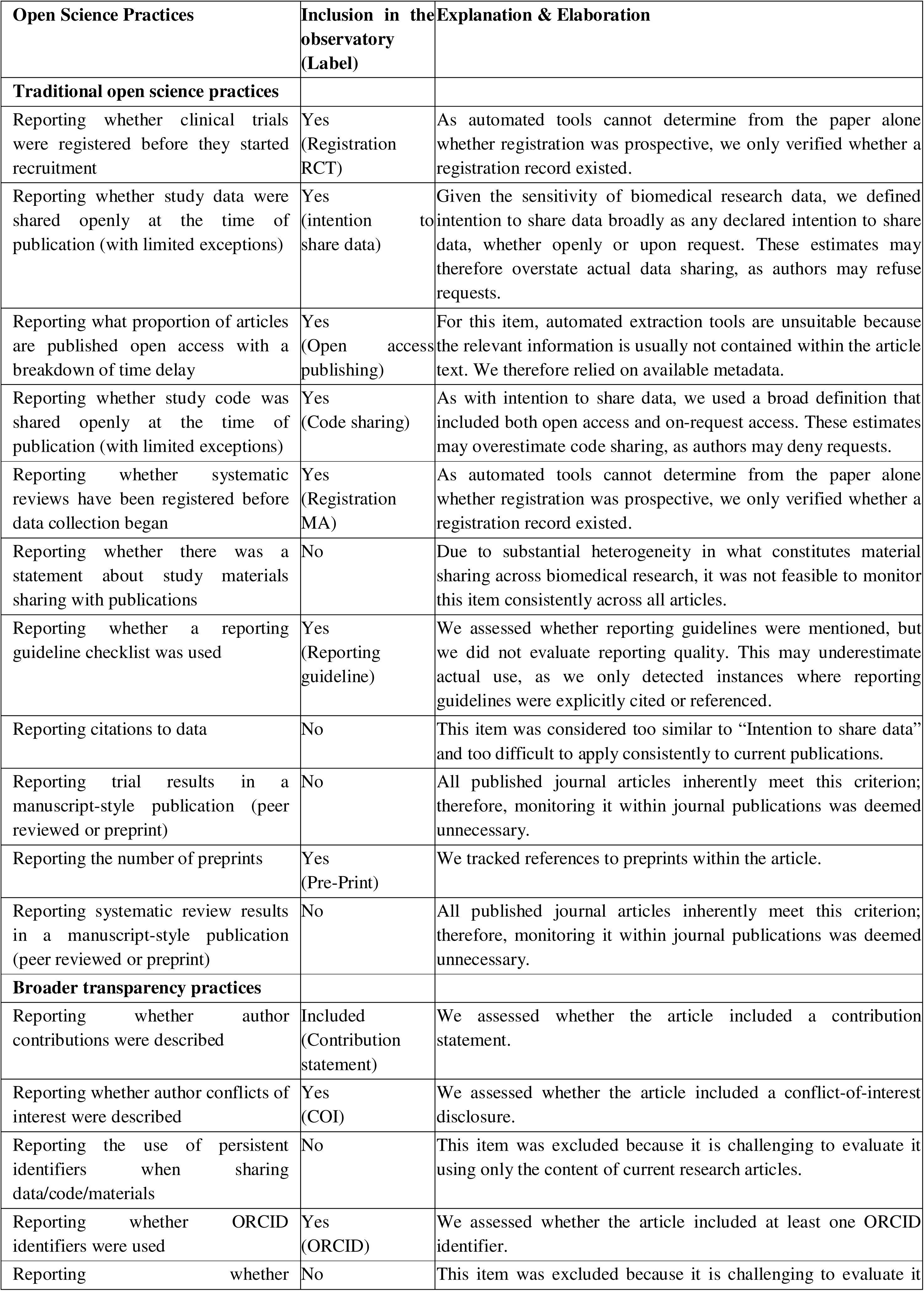
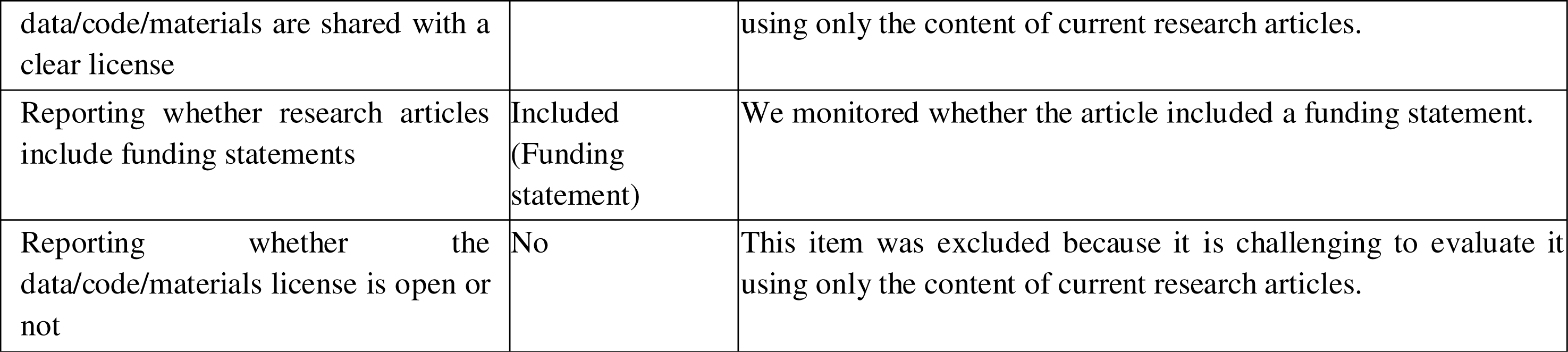
Description of the 12 Traditional Open Science practices and the 7 Broader transparency practices obtained from a Delphi study and decision to include it or not in the observatory(22).

#### Automated tools (index tests)

Leveraging our connections with the ScreenIT Working Group (21) and experts encountered through OSMI, we identified nine candidate tools that assess various aspects of these open science practices. For detailed technical specifications, please refer to **Web Appendix 4**. The tools we utilized comprise two proprietary solutions (SciScore (26) (RRID:SCR_016251) and DataSeer (27) (RRID:SCR_023027)), six open-access resources (rTransparent (28) (RRID:SCR_019276), ODDPub (29)(RRID:SCR_018385), ctregistries (30) (RRID:SCR_024412), ContriBOT (31) (RRID:SCR_027089), and TNRscreener (32)(RRID:SCR_019211), Unpaywall (33) (RRID:SCR_016471)), as well as an internally developed, fully local AI workflow built around Ollama running Llama-3.3-70B to ensure no data leave our environment. For Large Language Model (LLM) analysis, PDFs were first parsed to TEI/XML with Grobid; the XML was then split into manageable chunks and queried via LlamaIndex using standardized prompts that cover 14 transparency criteria, with strict boolean/categorical mappings to normalize outputs across articles. LLM behavior was validated on a dataset distinct from the one used in this study, with full details provided online (34). Where native converters exist (e.g., ODDPub, rTransparent), we relied on their Poppler-based pdf_to_text functions (35); otherwise we used Grobid (via a Python client) for PDF to XML conversion, and executed the end-to-end pipeline through containerized jobs (Docker to Apptainer) for scale and reproducibility.

#### Validation database

To evaluate and compare the diagnostic accuracy (ability to correctly identify the presence or absence of a specific open science practice) of these tools and select the most suitable one for each open science practice, a random sample of 300 research articles published between 2022 and 2023 was used, with equal distribution across journals and article types. If a journal did not publish a specific type of study, additional articles were randomly selected from other journals to ensure at least 100 articles per research article type. When an incorrect classification occurred due to PubMed extraction, the article was replaced with another randomly sampled one.

Manual extraction served as the reference standard (the best available benchmark for determining the true presence or absence of an open science practice, against which tools are compared) for assessing the diagnostic accuracy of each tool in detecting open science practices, consistent with most prior meta-research (28). Two examiners (FJA and MS) independently extracted data from the PDF and supplementary files using a pre-developed data extraction sheet. Any disagreements were resolved by a third examiner (CV). Detailed categorizations are provided in the data extraction file available on OSF: 10.17605/OSF.IO/F2VW9.

#### Statistical analysis

A descriptive analysis was conducted on articles that were manually extracted from the validation database. The open science practices of each journal, along with the results for various indicators, were summarized using frequencies and percentages. Weighted frequencies and the corresponding 95% confidence intervals were calculated for each open science practice, using the proportions from each journal and article category. This allowed correcting imbalances that were not addressed by our randomization process.

For each tool, we estimated how accurately it could detect each open science practice using three indicators: sensitivity (the ability of a tool to correctly identify the presence of an open science practice), specificity (the ability of a tool to correctly identify the absence of an open science practice), and the F1 score, along with their 95% confidence intervals. The F1 score is a single number that balances sensitivity and precision (how often the tool is correct when it says a practice is present) (36). It is especially useful to avoid both missing real cases and making too many false detections. For each practice, we selected the best tool based on its accuracy, ease of use, and whether it was openly available. These selected tools were then used to monitor open science practices in the chosen journals.

In addition, we performed exploratory analyses to assess each tool’s accuracy by journal and research type, aiming to identify formatting-related issues rather than compare journals.

#### Comparison between PDF and XML files

PDF files require conversion to text (XML files), a step that can lead to information loss (e.g., in tables). To assess its impact, we compared tool performance on PDF and XML inputs. The comparison used XML files extracted from Europe Pubmed Central (ePMC) (37).

### Observatory

#### Research articles 2020-2023

We tracked open science practices in all original research articles published in each selected journal, from 2020 to 2023. Those articles are included in our observatory.

#### Pre-post changes following the 2005 registration policy

We analyzed changes in registration practices before and after the 2005 ICMJE policy by tracking all RCTs published from 2003 to 2010, i.e. starting two years before and ending five years after the policy’s introduction. Given that our previous validation covered a shorter period and uptake of open science practices may change over time, we further assessed diagnostic accuracy using 150 randomly selected RCTs (15 from each journal) within this timeframe. Two independent reviewers (CV and APMD or MS) extracted data, resolving disagreements with FN. This process also helped estimate the rate of false positive RCTs in our search results.

#### Pre-post changes following data sharing policies

To assess the impact of different data sharing policies, we monitored intention to share data practices in all available RCTs (see Introduction and **Web Appendix 5** for a timeline). We focused on pre-post comparisons, covering two years before and after the policy change for each journal. For Annals of Internal Medicine, we included articles from 01/04/2005 to 01/04/2009; for PLOS Medicine, from 01/04/2012 to 01/04/2016. Given the more gradual changes at the BMJ, we monitored a longer period (25/03/2007 to 01/07/2017). For other journals, we focused on 2016–2020, surrounding the 2018 ICMJE policy. As with trial registration, we reassessed diagnostic accuracy using all articles for Annals of Internal Medicine, the BMJ and PLOS Medicine and 50 randomly selected from other journals. Data sharing practices were manually extracted (by CV and APMD), with disagreements resolved by FN. This process also helped estimate the rate of false positive RCTs in our search results.

#### Open Science practices

The protocol was registered before the start of any analysis on the Open Science Framework (OSF, https://doi.org/10.17605/OSF.IO/F2VW9) on 1 June 2024.

We used the STARD 2015 reporting guideline (38). Data and statistical code are openly available on the OSF: https://doi.org/10.17605/OSF.IO/F2VW9. The code is provided in R language and was run using R Project for Statistical Computing (RRID:SCR_001905) version 4.2.2, with the libraries tidyverse (RRID:SCR_019186) (39) and Patchwork (RRID:SCR_000072) for the data visualization . All analyses were performed on a MacBook Pro with an M3 Pro chip and on the Eskemm Data calculation server (40).

### Protocol modifications

#### Data extraction

Due to delays in obtaining protocol approval from all co-authors, data extraction started prior to registration (on 29^th^ April 2024). Importantly, no analyses were conducted before protocol registration, only manual extraction.

#### TOP 2025

We initially planned to evaluate journal policies using TOP2015 (41) but updated this to TOP2025 after the preprint was posted (20). As such, since no TOP assessments for the new criteria had been conducted yet, we did not use any assessments conducted by the Center for Open Science.

#### CoARA

We also planned to extract whether the journals/publishers supported the Coalition for Advancing Research Assessment (CoARA) but learned that journals/publishers could not endorse CoARA as they are not directly involved in research assessment.

#### Comparison XML PDF

In the audit, we report overall results as planned as well as findings for clinical trials, specifically, as some policies are limited to clinical trials. A sensitivity analysis comparing tool performance between XML and PDF files was initially planned on all articles from the diagnostic base instead of the ePMC subset. As not all articles are available on ePMC, this analysis was conducted only on those openly accessible on ePMC using the R package (42).

#### Weighted frequencies

Although not pre-specified in the protocol, weighted frequencies were applied to account for variability in study type and journal within our validation database.

#### Manual extraction of data sharing

Initially, we planned to assess policy effects on data sharing using only automated tools, but we also manually reviewed BMJ, PLOS Medicine, and Annals of Internal Medicine to verify ground truth and compare results with automated extraction.

#### Additional tool

We initially planned to assess all open science practices directly from the manuscript. However, because Open Access status cannot be determined from the manuscript itself and is generally obtained from article metadata, we evaluated this outcome using Unpaywall via the roadoi package (43).

### Patient and public involvement

We had no established contacts with specific patient groups who might be involved in this project. No patients were involved in setting the research question or the outcome measures, nor were they involved in the design and implementation of the study. There are no plans to involve patients in the dissemination of results, nor will we disseminate results directly to patients.

## Results

### TOP2025 journal policy audit

We collected TOP2025 data from ten journals. After outreach, five journals (The BMJ, CMAJ, JAMA, The Lancet, PLOS Medicine) responded, verified our data extraction, and 11 suggested amendments (some journals suggested several) that resulted in 6 edits to our ratings.

Figure 1 presents an overview of each journal or journal assessment in accordance with TOP 2025, overall, for all types of research articles and specifically about randomized controlled trials (RCTs). TOP factors ranged from 1 (New England Journal of Medicine) to 13 (PLoS Medicine). Many journals’ policies (i.e. registration, protocol sharing, statistical analysis plan sharing, data sharing and reporting transparency) applied specifically to clinical trials rather than to all research articles. No journals have clear policies on verification practices and only few on promoting verification studies, except Nature Medicine and PLoS Medicine, which supports replication studies, and BMC Medicine, which offers registered reports. In addition, of the 10 journals, 2 have signed the DORA declaration themselves, while 5 are covered through their publisher’s endorsement (including the 2 direct signatories). Full data and more details are shared on OSF 10.17605/OSF.IO/F2VW9.

**Figure 1:**
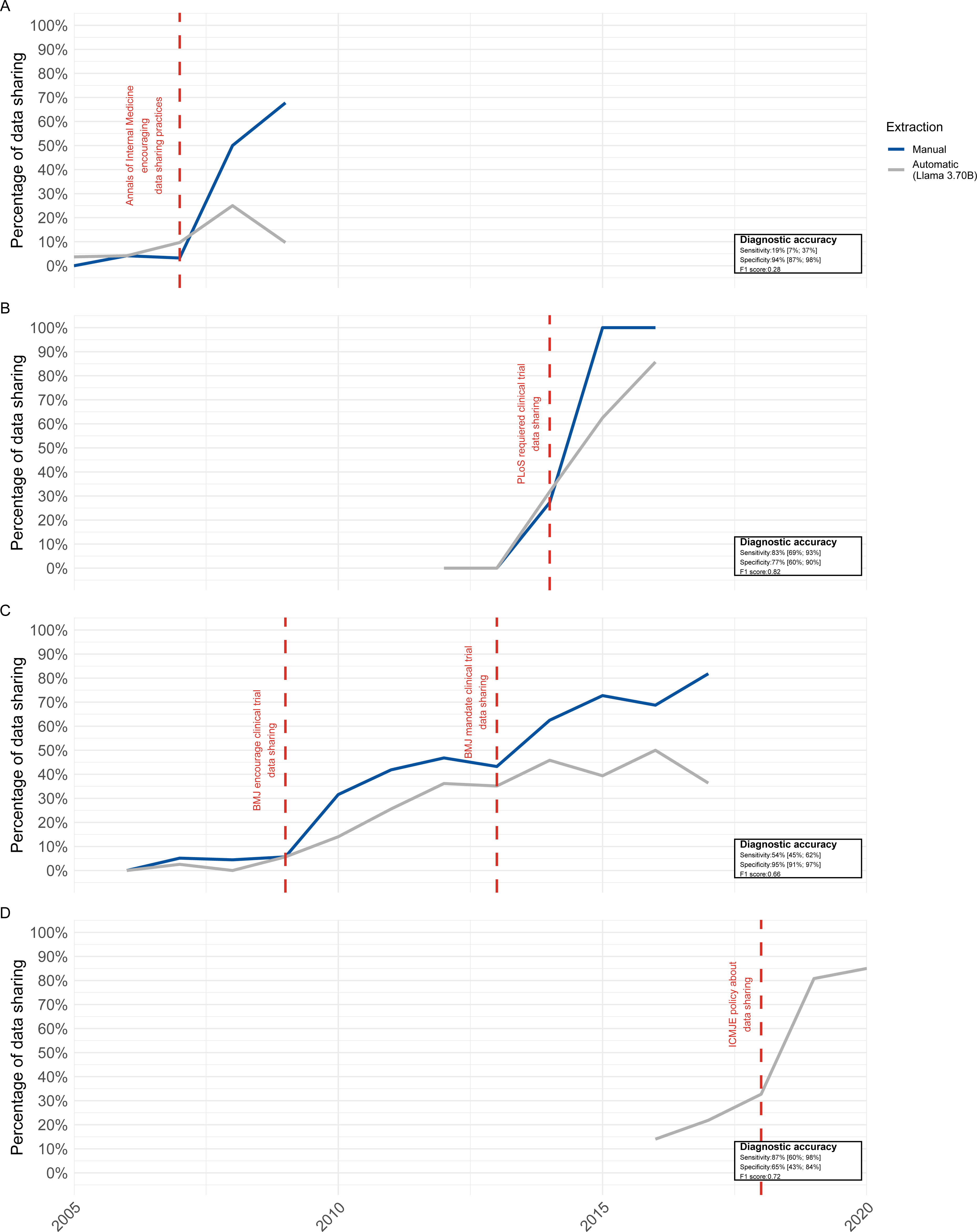
Figure Policy audit (TOP 2025) of 10 biomedical journals. *Panel A- indicate the score (0-3) for all study type. Panel B- indicate the score (0-3) for clinical trials* *Colors indicate scores: grey (0), light blue (1), medium bleu (2), dark blue (3). Bar plots show overall score distributions*.

### Research articles selection

PubMed searches were performed on 29^th^ April 2024. **Web Appendix 6** presents the study flow chart, outlining the selection process for the validation databases - detailing both included articles and those that were misclassified - as well as the observatory. Because of misclassifications related to study type, several articles were either excluded or reclassified, which required conducting multiple randomization rounds. In total, 312 research articles were included in the validation database (103 RCT, 98 MA and 111 other types of research article). Additional validation databases were extracted for registration and data sharing with details provided in **Web Appendix 7-8**.

### Diagnostic accuracy of tools to monitor open science practices

Not all journals published every type of research article. For instance, we found no meta-analysis published in *NEJM*. **Web-Appendix 9** presents the descriptive analysis of the validation database. Regarding transparency in reporting, conflicts of interest were disclosed in 100% of the research articles. Mention of preprints was rare, with only one identified in the validation database. Registration (98% vs 69% vs 18%), protocol sharing (89% vs 67% vs 23%), and statistical analysis plan sharing (51% vs 4% vs 8%) were more common in RCTs than in meta-analyses or other types of study. Authors most often indicated that data were “available upon request”, regardless of study type. Figure 2 presents the weighted frequencies for each open science practice based on the number of articles per journal and study type.

**Figure 2:**
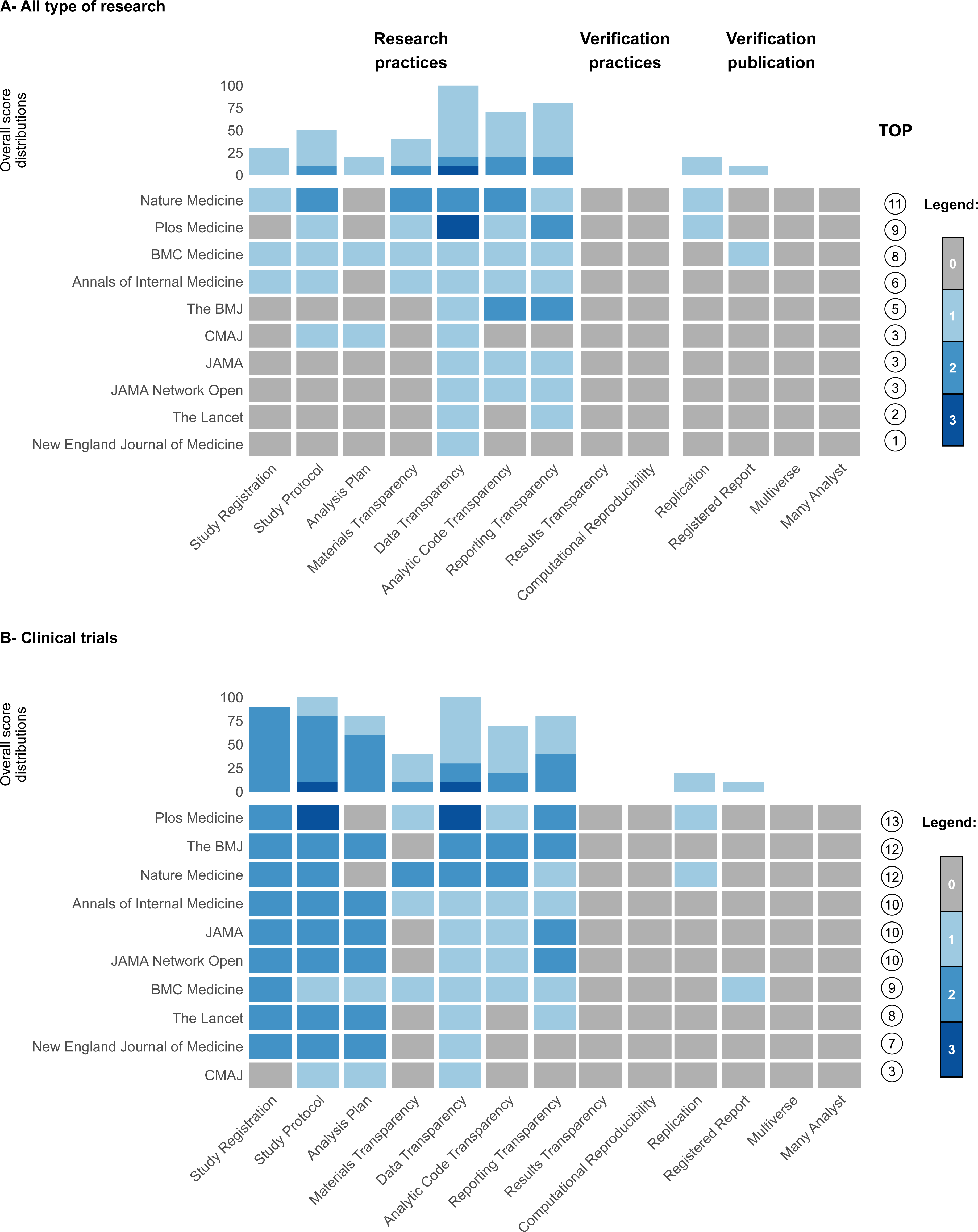
Observatory results: *Panel A-Tools: The selected tool based on its diagnostic performance and free of using* *Panel B-- A grey-to-blue gradient indicates tool performance, with darker blue representing better results. (NA: Not Applicable)* *Panel C- Percentages (with confidence intervals) of open science practices, adjusted for the number of articles published by study type and journal in the random sample. The orange around indicates the observed percentage in the global observatory obtained from the selected tools*.

The accuracy of automated tools varied depending on the practice evaluated, with F1-scores ranging from 0.06 (ORCID, Llama 3.70B) to 1.00 (Conflict of Interest statement, rTransparent). Comprehensive details for each tool are provided in **Web Appendix 10**.

Based on the performance and accessibility (when performance were similar), we retained for the observatory rTransparent for study registration (sensitivity=77% [70%-83%], specificity=93% [88%-97%]), Conflict of Interest statement (sensitivity=100% [99%-100%], specificity is uncalculable as all article have a statement) and funding statement (sensitivity=99% [97%-100%], specificity=67% [22%-96%]). ContriBOT was selected for author contributions (sensitivity=87% [82%-90%], specificity =78% [56%-93%]) and for the detection of ORCID number (sensitivity=86 % [79%-91%], specificity=99 % [96%-100%]). Unpaywall was selected for Open access publications (sensitivity=99% [96%-100%], specificity=36% [11%-69%]). For all other practices, we selected Llama 3.3 70B that frequently demonstrated the best performance, although F1 scores ranged from 0.06 to 1. Different diagnostic accuracies were observed depending on the type of research and the journal, likely due to different formatting standards (**Web Appendix 11-24**). Comprehensive information regarding all chosen tools, along with justifications for each open science practice, is presented in **Web Appendix 25.**

### Comparison of performance of selected automatic tools using PDF of XML files

This analysis to compare the efficacy of the tools using PDF or XML files focused on 182 of the 312 articles (58%) from the diagnostic base. For rTransparent, performance was better when registration data were extracted from PDF files. In contrast, file type had no impact on the detection of COI and funding statements. For ContriBOT, we observed the same results. Finally, for the analyses based on Llama 3.7B, the results varied considerably depending on the outcome. Overall, accuracies were similar, with a slight advantage when using XML files (except for the funding statement and randomization). The impact of file type on tool performance is described in **WebAppendix** 26.

### Observatory

#### Research articles 2020-2023

All open science practices identified in all 15,624 research articles published between 2020 and 2023 are presented in Figure 2. At the article level, available estimates indicate that the implementation of open science practices remained limited during that period. For instance, for registration, the overall estimate was 20% of articles, with variation observed across research types: 70% for randomized controlled trials (RCTs), 53% for meta-analyses, and 7.6% for other research articles. Automated tools tended to underestimate the adoption of most practices, except for protocol sharing, statistical analysis plan sharing, code sharing, and preprinting, which were overestimated.

#### Pre-post changes following the 2005 registration policy

For this longitudinal analysis, additional validation showed a sensitivity of 53% [42%; 64%], specificity of 94% [85%; 98%], and an F1 score of 0.67. 150 out of 167 studies (90%) in the validation sample were RCTs. Figure 3 shows registration practices for all studies identified as RCTs published between 2003 and 2010, before and after the 2005 ICMJE policy. In 2003, almost no RCTs were registered, whereas by 2010 estimates suggest a proportion of nearly 30%, with a marked increase after 2005.

**Figure 3:**
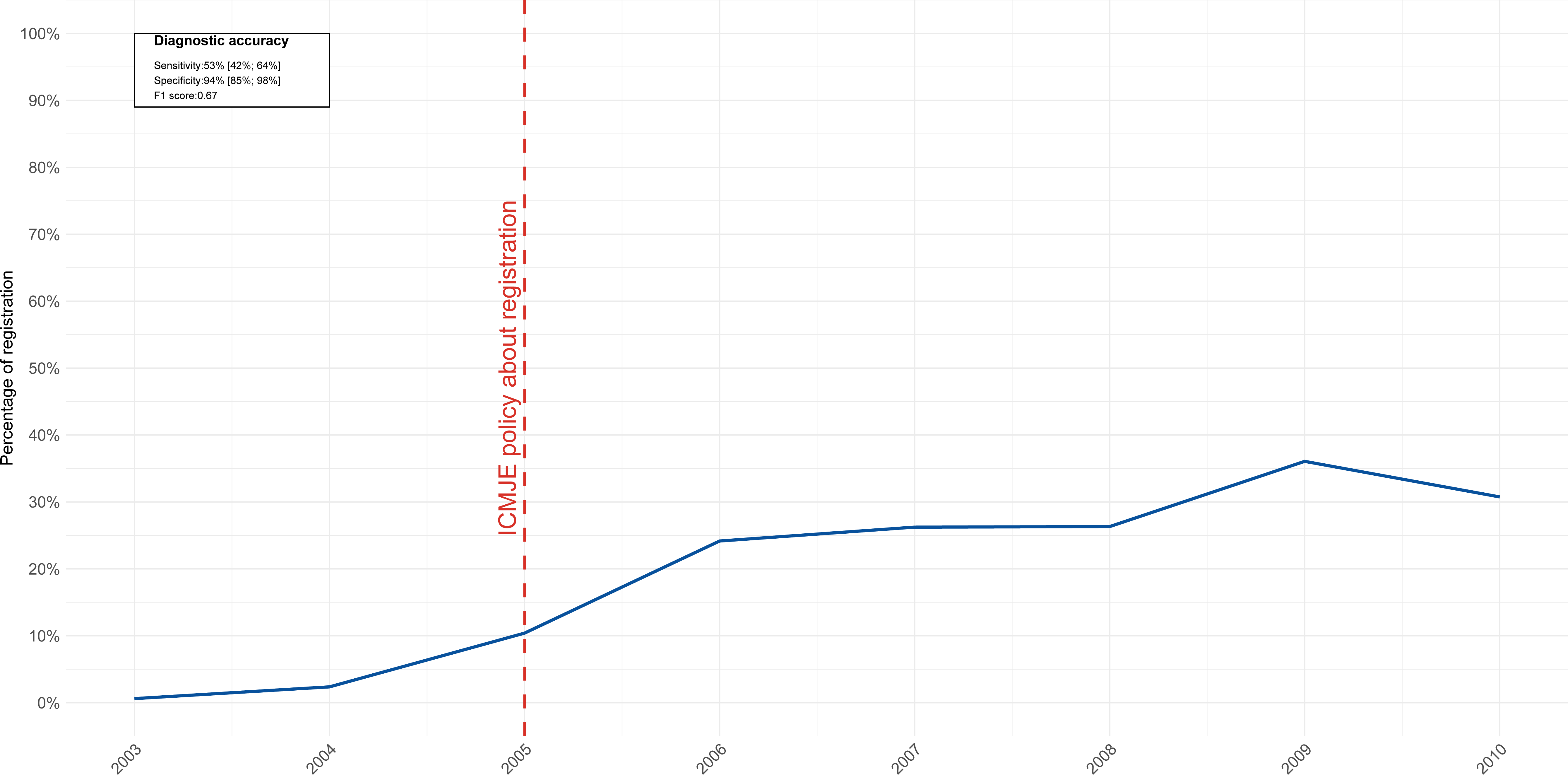
Evolution of registration practices for all RCTs published between 2003 and 2010, before and after the 2005 ICMJE policy.

#### Pre-post changes following data sharing policies

Manual extraction identified intention to share data in 140 out of 410 (34%) RCTs published in the BMJ between 2007 and 2017 (with 138/410 (34%) upon request and 2/410 (0.5%) openly available), in 43/79 RCTs (54%) published in PLOS medicine between 2012 and 2017 (with 15/79 (19%) upon request and 28/79 (35%) openly available) and in 31/129 papers (24%) in the Annals of Internal Medicine between 2005-2009 (all upon request). For the sample of RCTs from other journals, 22/50 (44%) have an intention to share data between 2016 and 2020 (with 21/50 (42%) upon request and 1/50 (2%) openly available). In the articles that were manually analyzed, 666 of 730 (91%) of the retrieved studies were RCTs.

Figure 4 represents intention to share data practices for all available RCTs across timelines aligned. In the BMJ, there was a sharp increase from 10% to 70% within a year following the introduction of a policy encouraging clinical trial data sharing. For PLOS Medicine and the global ICMJE policy, a similar upward trend in intention to share data was observed over the years. Manual extraction from Annals of Internal Medicine, The BMJ, and PLoS Medicine demonstrated that automated extraction tends to underestimate practices, though it accurately identifies overall trends.

**Figure 4:**
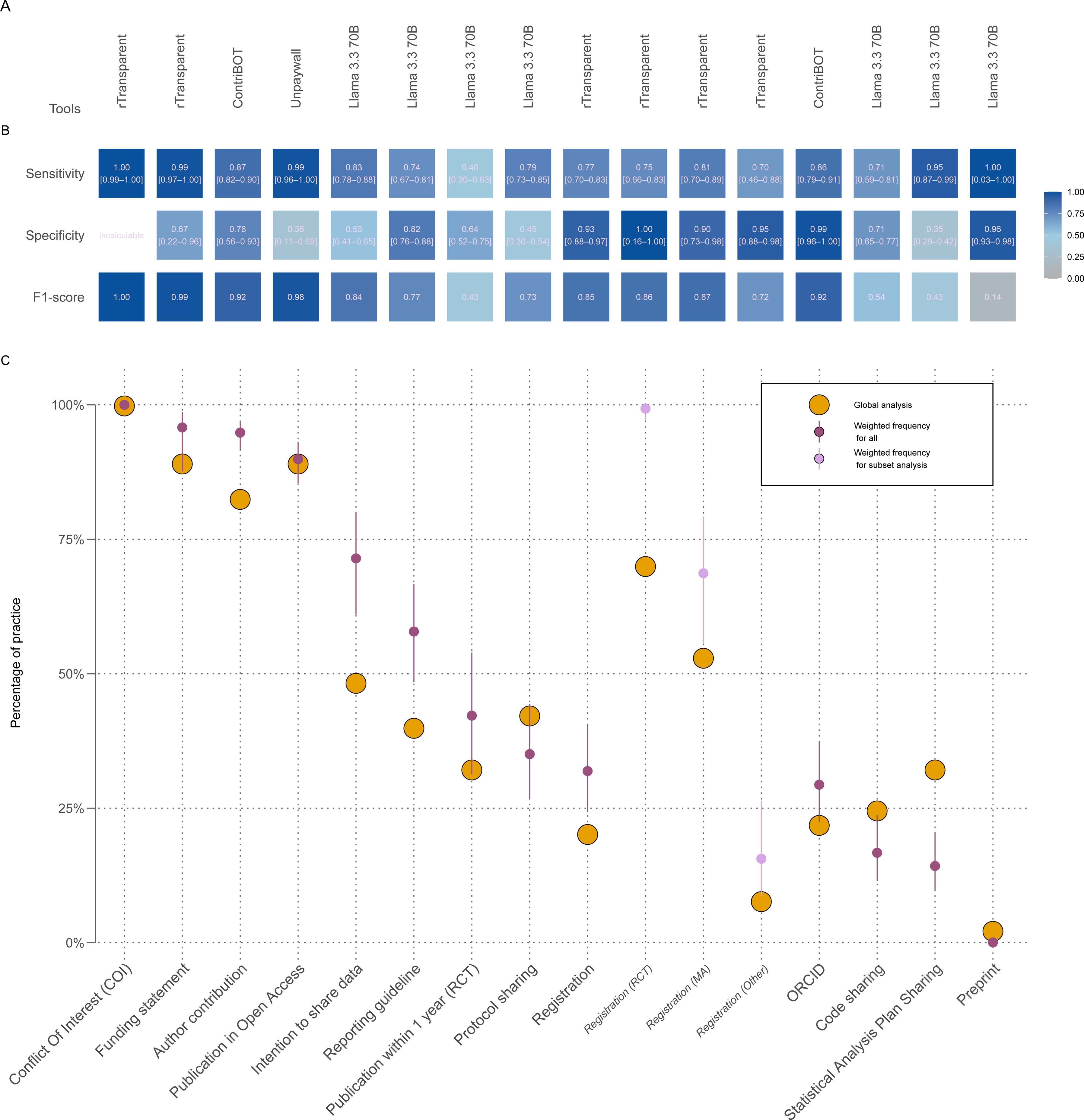
Evolution of the intention to share data practices for all RCTs. Panel A representation of Annals of internal Medicine intention to share data practice from 2005 to 2009 after the 2007 policy Panel B representation of PLoS Medicine intention to share data practice from 2012 to 2016 after the 2014 policy Panel C representation of The BMJ intention to share data practice from 2007 to 2017 after the 2009 and 2013 policies Panel D representation of other journals (except AIM, The BMJ and PLoS Medicine) intention to share data practice from 2016 to 2021 after the 2018 policy The blue line indicates the extraction realized by the automatic tool (Llama 3.70B) and the grey one indicates the manual extraction.

## Discussion

### Statement of principal findings

Among ten leading generalist medical journals, most had policies that aligned, at least partially, with the transparency standards set by TOP2025 (20). This alignment was particularly noticeable for policies concerning randomized controlled trials (RCTs), indicating that the transparency of RCT-related content receives more attention, compared to other study types. Overall, stronger policies are needed, particularly for non-RCTs and for practices such as registration, protocol sharing, statistical analysis plan sharing, and/or code sharing. Journal policies must also include guidance and requirements for verification practices and verification studies. Policies promoting verification practices of studies such as replication or registered reports are indeed rare. BMC Medicine was the only journal in our sample to explicitly offer registered reports. Although this format is gaining recognition in the broader scientific literature and has been adopted by journals such as Nature and PLOS Biology, its uptake remains very limited in generalist medical journals, including PLOS Medicine from the same publisher as PLOS Biology. On a positive note, we observed meaningful improvements in open science practices, such as registration and intention to share data, following policy changes, suggesting that these policies can drive real change. This should be interpreted with caution: we monitored intent to share data, not actual data sharing, which is especially complex for sensitive health data. As previously shown at BMJ, intent to share does not always result in true data sharing (44).

At the article level, our results were consistent with the overall trends observed in journal-level policy evaluations. Open science practices such as trial registration and data sharing appeared more frequently in randomized controlled trials (RCTs), which aligns with their prominent role in evidence-based medicine. Furthermore, obtaining patient consent for data sharing is typically more straightforward during RCT planning, making this practice more feasible than in studies using routinely collected clinical data. Another study design for which data sharing should be more straightforward, are meta-analyses using aggregated and already published data. Although data sharing poses minimal risk in these designs (45), data sharing remained uncommon.

### Strengths and weaknesses of the study

This study has several limitations. The diagnostic accuracy of several tools was below optimal levels, so all observatory indicators need to be interpreted carefully, keeping in mind the performance limitations highlighted in the validation database. Nevertheless, triangulating data from the policy audit and both manual and automated extraction offers consistent evidence for interpreting the results. It is also important to note that, for data sharing, tools such as Oddpub and rtransparent have been developed to detect open data (even if they also flag data-sharing upon request). In this study, we focused on intention to share, and using these tools had lower performance than the LLM; in fact, their performance was even better when evaluating intention to share than when detecting open data sharing alone. Then, although we used a carefully defined reference standard for data extraction, it remains an imperfect measure of actual journal practices. For example, few studies explicitly mention of reporting checklists (i.e. CONSORT (46)) in RCT articles, even though adherence to such standards is expected to be common practice. It is possible that checklists were used but not cited or attached, leading to underestimation of uptake by our data extractors. This issue is less likely to affect other open science practices but nonetheless highlights the limitations of relying solely on textual mentions to assess compliance. Third, rTransparent detects trial registration numbers reported in articles (e.g., NCT identifiers for clinical trials). As NCT numbers only emerged in 2000, the low percentage observed in early articles may partly reflect a lack of standardization at this time. And indeed, we analysed pre-post changes over a relatively short time window before and after policy changes. Larger effects may emerge over a longer period, although the tools were able to detect changes within the observed timeframe. Another technical limitation concerns the availability of supplementary materials. We restricted our analysis to materials available in PDF format, which prevented us from including supplements provided in other formats, such as Excel or Word files. In addition, PDF supplements consisting of images or scanned documents could not be parsed by the tools used in this study. This may have resulted in missed information for some articles. Moreover, both commercial tools (SciScore and DataSeer) only worked on a subset of the articles excluding, notably, the XML-formated articles from the Lancet. Given that limitations are intrinsic to all automated extraction tools, we recommend that all ongoing observatory initiatives share their diagnostic performance outcomes and related data to improve reproducibility.

Despite these challenges, the study has notable strengths. The tools used here, although imperfect, have been developed through rigorous processes and previously validated in the literature (28)(29)(47)(33)(48)(49). When applied to a large and diverse sample of journal articles, these tools can provide valuable insights into the evolution of transparency practices over time. Our study shows that these tools are particularly useful for tracking large-scale trends in journal policies, a feature that is crucial for informing evidence-based policy making. Last, the dataset included a variety of study types and provided an overview of transparency standards as implemented in leading journals.

### Strengths and weaknesses in relation to other studies, discussing important differences in results

In comparison to an earlier study using rTransparent, that mainly assessed Open Access articles from PubMed Central (28), the diagnostic accuracy of automated tools was lower when implemented on those journals, some of which include subscription-based content. This may be due to access restrictions, which prevented us from retrieving a complete dataset for all journals (e.g. the Lancet). Additionally, different journals vary greatly in how they present transparency-related information, with some including it in the main article and others placing it in supplementary materials. This variability may lead to discrepancies in tool outputs that reflect formatting artifacts rather than genuine differences in practice uptake. These limitations underscore the need for caution when using automated tools to compare journals. We therefore used tools descriptively to highlight general trends rather than to make normative comparisons or rankings.

### Meaning of the study: possible explanations and implications for clinicians and policymakers

Our findings demonstrate the potential value of independent monitoring systems for journal transparency practices. The indicators we used (drawn from the TOP2025 framework (20) and core open science practices (22)) are the result of structured expert consensus and provide a reliable basis for assessing journal performance. This type of monitoring could help authors to identify journals that align with their values, particularly those committed to openness and reproducibility. It could also offer research-performing organizations, including universities that have endorsed CoARA(50),an additional way to evaluate journal quality in line with responsible research evaluation principles. Moreover, systematic monitoring may help to distinguish legitimate journals that implement best practices from predatory journals that do not (24).

The development of such observatories may ultimately support evidence-informed policymaking and strengthen the open science infrastructure within biomedical research. However, these benefits will only be realized if technical challenges are addressed, particularly those related to formatting inconsistencies across journals. Current tools are valuable for identifying overall trends, but all tools have performance limitations, and tool results are best interpreted by a knowledgeable reader. Reported performance conflates multiple sources of error: conversion errors from input formats (such as PDF to text or XML processing), mistakes made by the tools themselves, and missing information in supplements that tools cannot easily access. While proposed solutions often target only one or two of these issues, improving input quality through better conversion or standardized journal layouts could enhance overall tool performance.

LLMs are expected to provide a generic, journal-format-agnostic solution capable of extracting multiple kinds of information without relying on ad hoc heuristics or specific procedures that require large engineering effort. Until such comprehensive capabilities are fully realized, the right combination of specialized tools together with possible improvements of LLM-based extraction (through, e.g., larger, multi-model LLM, or better prompting to extract information) means the implementation of such an observatory could become possible even at the article level. It is also important to ensure the responsible use of models, preferably locally to avoid data sharing with companies or require them to adopt policies against the misuse of shared data when accessed. There are also serious tradeoffs to using LLMs (e.g. environmental and energy costs, financial costs, computational intensiveness, time needed to run large datasets, need to control the model or do regular tests to assess performance in the case of changes, etc.), especially when simpler algorithms give comparable results (49).

### Unanswered questions and future research

Our results could inform long term scalability in journal monitoring or integration into global monitoring frameworks like UNESCO or TOP2025. However, further work is required to identify effective methods for structuring and formatting articles to facilitate automated assessment. Collaboration among stakeholders -including journals, publishers, funders, researchers, and monitoring initiatives- may be necessary to establish consensus on standardizing such information and determining its placement, whether as structured metadata, within the main article, or in supplementary materials.

While some publishers, such as PLOS, have begun to adopt monitoring practices (51), this should become a broader effort. The ICMJE could play a critical role in facilitating such collaboration. Ultimately, these efforts will help foster a more transparent, trustworthy, and reproducible biomedical research ecosystem.

## Data Availability

All data and code used to realize that study are openly shared on OSF: https://doi.org/10.17605/OSF.IO/F2VW9.

## Acknowledgements

We would like to thank Xavier Chard-Hutchinson (XCH) from the library of the University of Rennes Medical School for validating the research algorithm. We also wish to thank Anita Bandrowski from SciScore for allowing us to run SciScore on our diagnostic dataset. We also would like to thank Maia Salholz-Hillel for her help in the use of TNR screener. And we thank the Eskemm data team for giving us access to their solution.

## Contributions and guarantor information

CV: Conceptualization, Methodology, Software, Validation, Formal analysis, Investigation, Data curation, Writing – original draft, Visualization, Supervision.

APMD: Conceptualization, Data curation, Writing – review & editing. GF: Investigation, Writing – review & editing.

MM: Investigation, Writing – review & editing.

MS: Data curation, Writing – review & editing.

FJA: Data curation, Writing – review & editing.

MA: Conceptualization, Investigation, Writing – review & editing.

VN: Conceptualization, Software, Writing – review & editing.

TLW: Conceptualization, Writing – review & editing.

NJD: Conceptualization, Writing – review & editing.

GD: Conceptualization, Writing – review & editing.

GG: Conceptualization, Writing – review & editing.

GLBL: Investigation, Writing – review & editing.

IS: Conceptualization, Writing – review & editing, Funding acquisition.

MMGL: Conceptualization, Writing – review & editing, Funding acquisition.

FN: Conceptualization, Methodology, Validation, Writing – original draft, Visualization,

Supervision, Project administration, Funding acquisition.

FN and CV are the guarantors of the work and accept full responsibility for the conduct of the study, the integrity of the data, and the decision to publish.

## Ethics approval

Any ethics approval was needed for this study.

## Data sharing statement

### Transparency

The lead author and the manuscript guarantor state that the manuscript is an honest, accurate, and transparent account of the study reported, that no important aspects of the study have been omitted, and that any divergences from the study as planned have been explained.

### Copyright for publication

The Corresponding Author has the right to grant on behalf of all authors, and does so, a worldwide license to the Publishers and its licensees in perpetuity, in all forms, formats and media (whether known now or created in the future), to i) publish, reproduce, distribute, display and store the Contribution, ii) translate the Contribution into other languages, create adaptations, reprints, include within collections and create summaries, extracts and/or, abstracts of the Contribution, iii) create any other derivative work based on the Contribution, iv) exploit all subsidiary rights in the Contribution, v) include electronic links from the Contribution to third party material wherever it may be located; and, vi) license any third party to do any or all of the above."

#### List of abbreviation

API: Application Programming Interface
BMC: BioMed Central
The BMJ: The British Medical Journal
CMAJ: Canadian Medical Association Journal
CoARA: Coalition for Advancing Research Assessment
COI: Conflict Of Interest
DORA: Declaration On Research Assessment
ICMJE: International Committee of Medical Journal Editors
JAMA: Journal of the American Medical Association
ORCID: Open Researcher and Contributor ID
OSMI: Open science Monitoring Initiative
PDF: Portable Document Format
PLOS: Public Library of Science
PRESS: Peer Review of Electronic literature Search Strategies
RCT: Randomized Controlled Trials
STARD: Standards for Reporting of Diagnostic Accuracy Studies
TEI: Text Encoding Initiative
TOP: Transparency and Openness Promotion
XML: Extensible Markup Language

## References

1. Munafò MR, Nosek BA, Bishop DVM, Button KS, Chambers CD, Percie du Sert N, et al. A manifesto for reproducible science. Nat Hum Behav. 2017 Jan 10;1(1):0021.

2. De Angelis C, Drazen JM, Frizelle FA, Haug C, Hoey J, Horton R, et al. Clinical trial registration: a statement from the International Committee of Medical Journal Editors. N Engl J Med. 2004 Sep 16;351(12):1250–1.

3. Laine C, Goodman SN, Griswold ME, Sox HC. Reproducible Research: Moving toward Research the Public Can Really Trust. Ann Intern Med. 2007 Mar 20;146(6):450–3.

4. Taichman DB, Sahni P, Pinborg A, Peiperl L, Laine C, James A, et al. Data sharing statements for clinical trials. BMJ. 2017 Jun 5;357:j2372.

5. Naudet F, Siebert M, Pellen C, Gaba J, Axfors C, Cristea I, et al. Medical journal requirements for clinical trial data sharing: Ripe for improvement. PLOS Med. 2021 Oct 25;18(10):e1003844.

6. Hamilton DG, Hong K, Fraser H, Rowhani-Farid A, Fidler F, Page MJ. Prevalence and predictors of data and code sharing in the medical and health sciences: systematic review with meta-analysis of individual participant data. BMJ. 2023 Jul 11;382:e075767.

7. Groves T, Godlee F. Open science and reproducible research. BMJ. 2012 Jun 26;344:e4383.

8. Godlee F, Groves T. The new BMJ policy on sharing data from drug and device trials. BMJ. 2012 Nov 20;345:e7888.

9. Loder E, Groves T. The BMJ requires data sharing on request for all trials. BMJ. 2015 May 7;350:h2373.

10. Loder E, Macdonald H, Bloom T, Abbasi K. Mandatory data and code sharing for research published by The BMJ. BMJ. 2024 Mar 5;384:q324.

11. Bloom T, Ganley E, Winker M. Data Access for the Open Access Literature: PLOS’s Data Policy. PLOS Med. 2014 Feb 25;11(2):e1001607.

12. Naudet F, Patel CJ, DeVito NJ, Goff GL, Cristea IA, Braillon A, et al. Improving the transparency and reliability of observational studies through registration. BMJ. 2024 Jan 9;384:e076123.

13. Chambers CD, Tzavella L. The past, present and future of Registered Reports. Nat Hum Behav. 2022 Jan;6(1):29–42.

14. Anthony N, Tisseaux A, Naudet F. Published registered reports are rare, limited to one journal group, and inadequate for randomized controlled trials in the clinical field. J Clin Epidemiol. 2023 Aug;160:61–70.

15. Curry S. Let’s move beyond the rhetoric: it’s time to change how we judge research. Nature. 2018 Feb 7;554(7691):147–147.

16. Moher D, Bouter L, Kleinert S, Glasziou P, Sham MH, Barbour V, et al. The Hong Kong Principles for assessing researchers: Fostering research integrity. PLOS Biol. 2020 Jul 16;18(7):e3000737.

17. ECCC - Environment and Climate Change Canada, Cloutier M, Dacos M, Ministère de l’enseignement supérieur et de la recherche. Report of the G7 Open Science - Research on Research Sub-Working Group [Internet]. Ministère de l’enseignement supérieur et de la recherche; 2023 May [cited 2025 Nov 25]. Available from: https://hal-lara.archives-ouvertes.fr/hal-04415049

18. Open science | UNESCO [Internet]. 2025 [cited 2025 Nov 25]. Available from: https://www.unesco.org/en/open-science

19. Bobrov E, Bracco L, Dacos M, Fressengeas N, Hrynaszkiewicz I, Iarkaeva A, et al. The Principles of Open Science Monitoring [Internet]. Open Science Monitoring Initiative; 2025 Jul [cited 2025 Nov 25]. Available from: https://zenodo.org/doi/10.5281/zenodo.15807480

20. Grant S, Corker KS, Mellor DT, Stewart SLK, Cashin AG, Lagisz M, et al. TOP 2025: An Update to the Transparency and Openness Promotion Guidelines [Internet]. 2025 [cited 2025 Nov 25]. Available from: https://osf.io/nmfs6_v2

21. Max-Delbrück-Centrum BI für GC und. Berliner Institut für Gesundheitsforschung - Charité und Max-Delbrück-Centrum. [cited 2025 Nov 25]. Automated screening tools - Service - BIH at Charité. Available from: https://www.bihealth.org/en/quest/service/service/automated-screening-tools

22. Cobey KD, Haustein S, Brehaut J, Dirnagl U, Franzen DL, Hemkens LG, et al. Community consensus on core open science practices to monitor in biomedicine. PLOS Biol. 2023 Jan 24;21(1):e3001949.

23. Laine C, Babski D, Bachelet VC, Bärnighausen TW, Baethge C, Bibbins-Domingo K, et al. Predatory journals: what can we do to protect their prey? BMJ. 2025 Jan 7;388:q2850.

24. Siebert M, Bourgeois FT, Naudet F. ICMJE Should Create a Certification System to Identify Predatory Journals. JAMA. 2025 Jul 1;334(1):87–8.

25. McGowan J, Sampson M, Salzwedel DM, Cogo E, Foerster V, Lefebvre C. PRESS Peer Review of Electronic Search Strategies: 2015 Guideline Statement. J Clin Epidemiol. 2016 Jul;75:40–6.

26. Menke J, Roelandse M, Ozyurt B, Martone M, Bandrowski A. The Rigor and Transparency Index Quality Metric for Assessing Biological and Medical Science Methods. iScience. 2020 Nov;23(11):101698.

27. DataSeer [Internet]. [cited 2025 Nov 25]. DataSeer. Available from: https://dataseer.ai/

28. Serghiou S, Contopoulos-Ioannidis DG, Boyack KW, Riedel N, Wallach JD, Ioannidis JPA. Assessment of transparency indicators across the biomedical literature: How open is open? PLOS Biol. 2021 Mar 1;19(3):e3001107.

29. Riedel N, Kip M, Bobrov E. ODDPub – a Text-Mining Algorithm to Detect Data Sharing in Biomedical Publications. Data Sci J [Internet]. 2020 Oct 29 [cited 2025 Nov 25];19(1). Available from: https://datascience.codata.org/articles/dsj-2020-042

30. GitHub [Internet]. [cited 2025 Nov 25]. ctregistries. Available from: https://github.com/maia-sh/ctregistries/commit/06c169cfa241ef8feda9e8b78f57b3012c85afd8

31. ContriBOT [Internet]. [cited 2025 Nov 25]. Available from: https://github.com/quest-bih/ContriBOT/

32. GitHub [Internet]. [cited 2025 Nov 25]. TRNscreener. Available from: https://github.com/bgcarlisle/TRNscreener/commit/9d02549a8d2ede3995013c8347aa33e0c9220427

33. Else H. How Unpaywall is transforming open science. Nature. 2018 Aug 15;560(7718):290–1.

34. Achar. GitLab. 2025 [cited 2025 Nov 25]. ollama. Available from: https://gitlab.inria.fr/glebartz/mr_oaso/-/tree/7c32d6ba0ef0e45af9ac3e73a9802122fa1fc788/ollama

35. Hornik K. Rpoppler: PDF Tools Based on Poppler [Internet]. 2013 [cited 2025 Nov 25]. p. 0.1-3. Available from: https://CRAN.R-project.org/package=Rpoppler

36. Acharya N. Understanding Precision, Recall, F1-score, and Support in Machine Learning Evaluation [Internet]. Medium. 2024 [cited 2025 Nov 25]. Available from: https://medium.com/@nirajan.acharya777/understanding-precision-recall-f1-score-and-support-in-machine-learning-evaluation-7ec935e8512e

37. Home - Europe PMC [Internet]. [cited 2025 Nov 25]. Available from: https://europepmc.org/

38. Cohen JF, Korevaar DA, Altman DG, Bruns DE, Gatsonis CA, Hooft L, et al. STARD 2015 guidelines for reporting diagnostic accuracy studies: explanation and elaboration. BMJ Open. 2016 Nov 14;6(11):e012799.

39. Wickham H, Averick M, Bryan J, Chang W, McGowan L, François R, et al. Welcome to the Tidyverse. J Open Source Softw. 2019 Nov 21;4(43):1686.

40. Eskemm Data [Internet]. eskemm. [cited 2025 Nov 25]. Available from: https://www.eskemm-numerique.fr/eskemm-data/

41. Nosek BA, Alter G, Banks GC, Borsboom D, Bowman SD, Breckler SJ, et al. Promoting an open research culture. Science. 2015 Jun 26;348(6242):1422–5.

42. Jahn N. europepmc: R Interface to the Europe PubMed Central RESTful Web Service [Internet]. 2016 [cited 2025 Nov 25]. p. 0.4.3. Available from: https://CRAN.R-project.org/package=europepmc

43. Jahn N. roadoi: Find Free Versions of Scholarly Publications via Unpaywall [Internet]. 2017 [cited 2025 Nov 25]. p. 0.7.3. Available from: https://CRAN.R-project.org/package=roadoi

44. Naudet F, Sakarovitch C, Janiaud P, Cristea I, Fanelli D, Moher D, et al. Data sharing and reanalysis of randomized controlled trials in leading biomedical journals with a full data sharing policy: survey of studies published in The BMJ and PLOS Medicine. BMJ. 2018 Feb 13;360:k400.

45. Cristea IA, Naudet F, Caquelin L. Meta-research studies should improve and evaluate their own data sharing practices. J Clin Epidemiol. 2022 Sep;149:183–9.

46. Hopewell S, Chan AW, Collins GS, Hróbjartsson A, Moher D, Schulz KF, et al. CONSORT 2025 statement: updated guideline for reporting randomised trials. BMJ. 2025 Apr 14;389:e081123.

47. Roelandse M, Ozyurt IB, Evanko D, Bandrowski A. Assessing the Effectiveness of SciScore in Supporting the Reproducibility of Scientific Research. Sci Ed. 46(2):46–52.

48. Iarkaeva A, Nachev V, Bobrov E. Workflow for detecting biomedical articles with underlying open and restricted-access datasets. PLOS ONE. 2024 May 8;19(5):e0302787.

49. Eckmann P, Barnett A, Bannach-Brown A, Atria EPB, Cabanac G, Franzen LDO, et al. Use as Directed? A Comparison of Software Tools Intended to Check Rigor and Transparency of Published Work [Internet]. arXiv; 2025 [cited 2025 Nov 25]. Available from: http://arxiv.org/abs/2507.17991

50. CoARA – Coalition for Advancing Research Assessment [Internet]. [cited 2025 Nov 25]. Available from: https://www.coara.org/

51. PLOS. Collaboratively seeking better solutions for monitoring Open Science [Internet]. The Official PLOS Blog. 2024 [cited 2025 Nov 25]. Available from: https://theplosblog.plos.org/2024/03/collaboratively-seeking-better-solutions-for-monitoring-open-science/

