## Supplements for "Monitoring of Open Science practices: a survey of 10 major medical journals"

**WEB APPENDIX**

**Web Appendix 1: List and description of the items evaluated by TOP 2025**

| **Research Practices** | | |  |
| --- | --- | --- | --- |
| **Practice** | **Level 1: Disclosed** | **Level 2: Shared and Cited** | **Level 3: Certified** |
| Study  Registration | Authors stated whether or not a study was registered—and, if so, where and when it was registered. | Researchers registered the study and cited the registration. | A party independent from the researchers certified that the study was registered at an appropriate time and the registration was complete per best-practice for the study design. |
| Study  Protocol | Authors stated whether or not the study protocol is available—and, if so, where and when it was shared. | Researchers publicly shared the study protocol and cited the protocol. | A party independent from the researchers certified that the study protocol was shared at an appropriate time and the study protocol was complete per best-practice for the study design. |
| Analysis  Plan | Authors stated whether or not the analysis plan is available—and, if so, where and when it was shared. | Researchers publicly shared the analysis plan and cited the analysis plan. | A party independent from the researchers certified that the analysis plan was shared at an appropriate time and the analysis plan was complete per best- practice for the study design. |
| Materials Transparency | Authors stated whether or not materials are available—and, if so, where. | Researchers cited materials deposited in a trusted repository by themselves or others. | A party independent from the researchers certified that materials were deposited and documented per best-practice for the type of materials. |
| Data  Transparency | Authors stated whether or not data are available—and, if so, where. | Researchers cited data deposited in a trusted repository by themselves or others. | A party independent from the researchers certified that data were deposited with metadata per best- practice for the type of data. |
| Analytic Code  Transparency | Authors stated whether or not analytic code is available—and, if so, where. | Researchers cited analytic code deposited in a trusted repository by themselves or others. | A party independent from the researchers certified that analytic code was deposited and documented per relevant best-practice. |
| Reporting Transparency | Authors stated whether or not they used a reporting guideline—and, if so, which guideline. | Authors publicly shared a completed reporting guideline checklist and cited the reporting guideline. | A party independent from the researchers certified that the researchers adhered to the appropriate reporting guideline for the study design. |

| **Verification Practices** | |
| --- | --- |
| **Practice** | **Definition** |
| Results Transparency | A party independent from the researchers verified that results have not been reported selectively based on the nature of the findings. To verify, the independent party can check that the study registration, protocol, and analysis plan match the final report–and the final report acknowledges any deviations. |
| Computational Reproducibility | A party independent from the researchers verified that reported results reproduce using the same data and following the same computational procedures. To verify, the independent party can check that they obtain the same results using data and code deposited in a trusted repository. |

| **Verification Studies** | |
| --- | --- |
| **Study Type** | **Definition** |
| Replication | A study that aims to provide diagnostic evidence about claims from a prior study by repeating the original study procedures in a new sample. |
| Registered Report | A registered study in which a study protocol and analysis plan are peer reviewed, and the study is pre-accepted by a publication outlet, before the research is undertaken. |
| Multiverse | A study in which a single research team examines the research question of interest across different, reasonable choices for processing and analyzing the same data. |

**Web Appendix 2:** Search strategies were reviewed by an information specialist

| Study type | Research algorithm |
| --- | --- |
| RCT | (2020:2023[Date - Publication] AND "hasabstract" AND randomizedcontrolledtrial[Filter] AND ("Annals of Internal Medicine"[Journal] OR "BMC Medicine"[Journal] OR "BMJ"[Journal] OR "CMAJ"[Journal] OR "JAMA"[Journal] OR "JAMA Netw Open"[Journal] OR "Lancet"[Journal] OR "Nat Med"[Journal] OR "N Engl J Med"[Journal] OR "PLOS Med"[Journal])) |
| Meta research | (2020:2023[Date - Publication] AND "hasabstract" AND meta-analysis[Filter] AND ("Annals of Internal Medicine"[Journal] OR "BMC Medicine"[Journal] OR "BMJ"[Journal] OR "CMAJ"[Journal] OR "JAMA"[Journal] OR "JAMA Netw Open"[Journal] OR "Lancet"[Journal] OR "Nat Med"[Journal] OR "N Engl J Med"[Journal] OR "PLOS Med"[Journal])) |
| Other research article | ((2020:2023[Date - Publication]) AND "hasabstract" NOT ("comment"[Publication Type]) NOT ("letter"[Publication Type]) NOT ("editorial"[Publication Type]) NOT ("published erratum"[Publication Type]) NOT ("news"[Publication Type]) NOT ("introductory journal article"[Publication Type]) NOT ("biography"[Publication Type]) NOT ("portrait"[Publication Type]) NOT ("congress"[Publication Type]) NOT ("interview"[Publication Type]) NOT ("retraction of publication"[Publication Type]) NOT ("personal narrative"[Publication Type]) NOT ("retracted publication"[Publication Type]) NOT ("patient education handout"[Publication Type]) NOT ("lecture"[Publication Type]) NOT ("autobiography"[Publication Type]) NOT ("clinical conference"[Publication Type]) NOT ("classical article"[Publication Type]) NOT ("address"[Publication Type]) NOT ("legal case"[Publication Type]) NOT ("expression of concern"[Publication Type]) NOT ("festschrift"[Publication Type]) NOT ("overall"[Publication Type]) NOT ("bibliography"[Publication Type]) NOT ("corrected and republished article"[Publication Type]) NOT ("interactive tutorial"[Publication Type]) NOT ("duplicate publication"[Publication Type]) NOT ("directory"[Publication Type]) NOT ("newspaper article"[Publication Type]) NOT ("periodical index"[Publication Type]) NOT ("dictionary"[Publication Type]) NOT ("meta analysis"[Publication Type]) NOT ("randomized controlled trial"[Publication Type])) AND ("Annals of Internal Medicine"[Journal] OR "BMC Medicine"[Journal] OR "BMJ"[Journal] OR "CMAJ"[Journal] OR "JAMA"[Journal] OR "JAMA Netw Open"[Journal] OR "Lancet"[Journal] OR "Nat Med"[Journal] OR "N Engl J Med"[Journal] OR "PLOS Med"[Journal]) |

**Web Appendix 3: Table of the available type of document for each journal**

| **Journal** | **PDF article** | **XML article** | **supplement** |
| --- | --- | --- | --- |
| Annals of Internal Medicine | X |  | X |
| BMC Medicine | X | X | X |
| The BMJ | X | X | X |
| CMAJ | X |  | X |
| JAMA | X |  | X |
| JAMA Network Open | X | X | X |
| The Lancet |  | X |  |
| Nature Medicine | X | X* | X |
| New England Journal of Medicine | X |  | X |
| Plos Medicine | X | X | X |

**Only the Open access articles of Nature Medicine are available*

**Web Appendix 4: table of the automatic tools with the practices that they assess**

| **Tool** | **Technical approach** | **RCT** | **Registration** | **Data sharing** | **Open access publishing** | **Code sharing** | **Publication 1year (for RCT*)** | **Protocol** | **SAP** | **Reporting Guideline** | **Preprint** | **Author contribution** | **COI** | **ORCID** | **Funding statement** |
| --- | --- | --- | --- | --- | --- | --- | --- | --- | --- | --- | --- | --- | --- | --- | --- |
| ODDPub^1^ | regex |  |  | **X** |  | **X** |  |  |  |  |  |  |  |  |  |
| rTransparent^2^ | regex |  | **X** | **X** |  | **X** |  |  |  |  |  |  | **X** |  | **X** |
| CTregistries^3^ | regex |  | **X** |  |  |  |  |  |  |  |  |  |  |  |  |
| TRN screener^4^ | regex |  | **X** |  |  |  |  |  |  |  |  |  |  |  |  |
| ContriBOT^5^ | regex |  |  |  |  |  |  |  |  |  |  | **X** |  | **X** |  |
| SciScore^6^ | regex | **X** | **X** | **X** |  | **X** |  |  |  |  |  |  |  |  |  |
| DataSeer^7^ | LLM + regex |  | **X** | **X** |  | **X** |  |  |  |  | **X** |  |  |  |  |
| Llama 3.3 70B ^8^ | LLM | **X** | **X** | **X** | **X** | **X** | **X** | **X** | **X** | **X** | **X** | **X** | **X** | **X** | **X** |
| Unpaywall^9^ | Web harvesting |  |  |  | **X** |  |  |  |  |  |  |  |  |  |  |

1- [*https://github.com/quest-bih/oddpub*](https://github.com/quest-bih/oddpub)
2- <https://github.com/serghiou/rtransparent>
3- [*https://github.com/quest-bih/ctregistries*](https://github.com/quest-bih/ctregistries)
4- [*https://github.com/bgcarlisle/TRNscreener*](https://github.com/bgcarlisle/TRNscreener)
5- [*https://github.com/quest-bih/ContriBOT*](https://github.com/quest-bih/ContriBOT)
6- [*https://sciscore.com/*](https://sciscore.com/)
7- [*https://dataseer.ai/*](https://dataseer.ai/)
8- [*https://gitlab.inria.fr/glebartz/mr_oaso/-/tree/dev-margaux/ollama?ref_type=heads*](https://gitlab.inria.fr/glebartz/mr_oaso/-/tree/dev-margaux/ollama?ref_type=heads)
9- [*https://unpaywall.org/*](https://unpaywall.org/)

*Abbreviations:*

*RCT: Randomized Control Trial (ability to detect if the tool is or not a randomized control trial).*

*SAP: Statistical Analysis Plan*

*COI: Conflict of Interest*

*ORCID: Open Researcher and Contributor ID*

| Journal | Practice | 2003 | 2004 | 2005 | 2006 | 2007 | 2008 | 2009 | 2010 | 2011 | 2012 | 2013 | 2014 | 2015 | 2016 | 2017 | 2018 | 2019 | 2020 | 2021 | 2022 | 2023 |
| --- | --- | --- | --- | --- | --- | --- | --- | --- | --- | --- | --- | --- | --- | --- | --- | --- | --- | --- | --- | --- | --- | --- |
| BMC med | Registration |  |  |  |  |  |  |  |  |  |  |  |  |  |  |  |  |  |  |  |  |  |
| CMAJ | Registration |  |  |  |  |  |  |  |  |  |  |  |  |  |  |  |  |  |  |  |  |  |
| JAMA | Registration |  |  |  |  |  |  |  |  |  |  |  |  |  |  |  |  |  |  |  |  |  |
| JAMA Network | Registration |  |  |  |  |  |  |  |  |  |  |  |  |  |  |  |  |  |  |  |  |  |
| Lancet | Registration |  |  |  |  |  |  |  |  |  |  |  |  |  |  |  |  |  |  |  |  |  |
| NEJM | Registration |  |  |  |  |  |  |  |  |  |  |  |  |  |  |  |  |  |  |  |  |  |
| AIM | Registration |  |  |  |  |  |  |  |  |  |  |  |  |  |  |  |  |  |  |  |  |  |
| BMJ | Registration |  |  |  |  |  |  |  |  |  |  |  |  |  |  |  |  |  |  |  |  |  |
| Plos Med | Registration |  |  |  |  |  |  |  |  |  |  |  |  |  |  |  |  |  |  |  |  |  |
| Nature | Registration |  |  |  |  |  |  |  |  |  |  |  |  |  |  |  |  |  |  |  |  |  |

**Web Appendix 5: Timeline the monitoring of registration (A) and data sharing (B) practices in randomized control trials for each journal, to assess the impact of the different policies.**

A- **For Registration**

To assess the impact of different registration policies, *we monitor changes in registration practices before and after the 2005 ICMJE policy by tracking all RCTs published from 2003 to 2010, i.e. starting two years before and ending five years after the policy's introduction/*

B- **Data sharing**

*To assess the impact of different data sharing policies, we monitored data sharing practices in all available RCTs. We focused on pre-post comparisons, covering two years before and after the policy change for each journal. 1- For Annals of Internal Medicine, we included articles from 01/04/2005 to 01/04/2009; 2- for PLOS Medicine, from 01/04/2012 to 01/04/2016. 3- the BMJ, we monitored a longer period (25/03/2007 to 01/07/2017) 4- For other journals, we focused on 2016–2020, surrounding the 2018 ICMJE policy.*

| Journal | Practice | 2003 | 2004 | 2005 | 2006 | 2007 | 2008 | 2009 | 2010 | 2011 | 2012 | 2013 | 2014 | 2015 | 2016 | 2017 | 2018 | 2019 | 2020 | 2021 | 2022 | 2023 |
| --- | --- | --- | --- | --- | --- | --- | --- | --- | --- | --- | --- | --- | --- | --- | --- | --- | --- | --- | --- | --- | --- | --- |
| BMC med | Data sharing |  |  |  |  |  |  |  |  |  |  |  |  |  |  |  |  |  |  |  |  |  |
| CMAJ | Data sharing |  |  |  |  |  |  |  |  |  |  |  |  |  |  |  |  |  |  |  |  |  |
| JAMA | Data sharing |  |  |  |  |  |  |  |  |  |  |  |  |  |  |  |  |  |  |  |  |  |
| JAMA Network | Data sharing |  |  |  |  |  |  |  |  |  |  |  |  |  |  |  |  |  |  |  |  |  |
| Lancet | Data sharing |  |  |  |  |  |  |  |  |  |  |  |  |  |  |  |  |  |  |  |  |  |
| NEJM | Data sharing |  |  |  |  |  |  |  |  |  |  |  |  |  |  |  |  |  |  |  |  |  |
| AIM | Data sharing |  |  |  |  |  |  |  |  |  |  |  |  |  |  |  |  |  |  |  |  |  |
| BMJ | Data sharing |  |  |  |  |  |  |  |  |  |  |  |  |  |  |  |  |  |  |  |  |  |
| Plos Med | Data sharing |  |  |  |  |  |  |  |  |  |  |  |  |  |  |  |  |  |  |  |  |  |
| Nature | Data sharing |  |  |  |  |  |  |  |  |  |  |  |  |  |  |  |  |  |  |  |  |  |

**Web Appendix 6: Study flow chart, outlining the selection process for the validation databases**.

*This figure detailing both included articles and those that were misclassified*

*Abbreviation: RCT: Randomized Control Trial, MA: Meta-analysis*

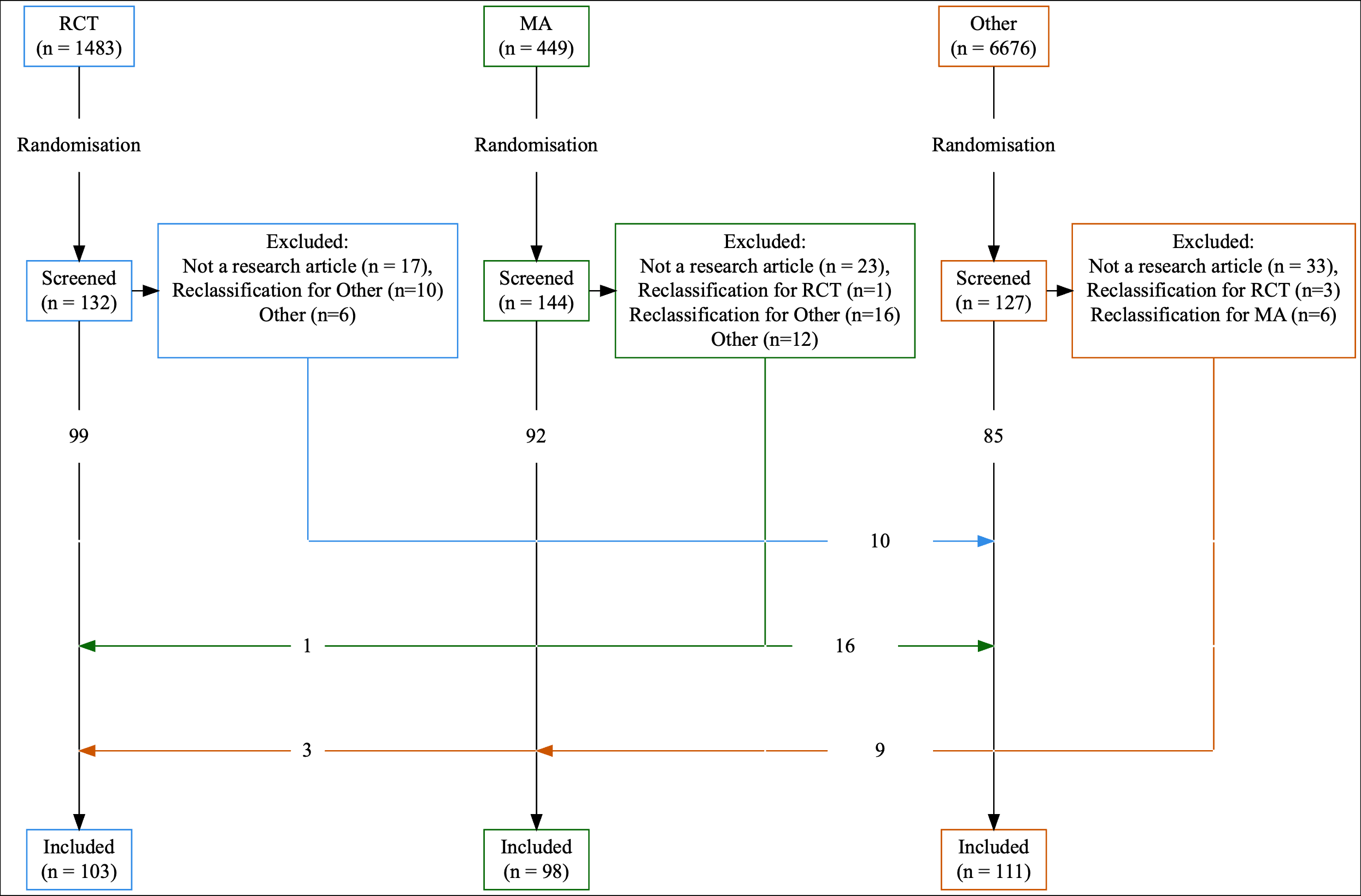

**Web Appendix 7: Manually extracted validation data for registration from 2003 to 2010**

| **Journal** | **Registration practice (2003-2010)** |
| --- | --- |
| AIM | 12/20 (60,00%) |
| BMC Med | 8/15 (53.33%) |
| BMJ | 12/21(57.14%) |
| CMAJ | 4/15 (26.67%) |
| JAMA | 12/21 (57.14%) |
| Lancet | 17/22 (77.27%) |
| NEJM | 10/18 (55.56%) |
| Nature Med | 0/3 (0,00%) |
| PLOS Med | 11/15 (73.33%) |
| **Overall** | 86/150(57.33%) |

**Web Appendix 8: Manually extracted validation data for data sharing from 2016 to 2020**

| **Journal** | **Data sharing (open access)** | **Data sharing (under request)** | **Data sharing (overall)** |
| --- | --- | --- | --- |
| BMC Med | 0/2 (0%) | 1/2 (50.00%) | 1/2 (50.00%) |
| CMAJ | 0/2 (0%) | 2/2 (100%) | 2/2 (100%) |
| JAMA | 0/11 (0%) | 5/11 (45.45%) | 5/11 (45.45%) |
| JAMA NWO | 0/5 (0%) | 1/5 (20.00%) | 1/5 (20.00%) |
| Lancet | 0/9 (0%) | 3/9 (33.33%) | 3/9 (33.33%) |
| NEJM | 1/21 (4,76%) | 9/21 (42.86%) | 10/21 (47.62 |
| **Overall** | 1/50 (2.00%) | 21/50 (42.00%) | 22/50 (44.00%) |

**Web Appendix 9***:* **Use of open science practices in the validation database obtained by manual extraction (considered as the gold standard).**

| **Characteristic** | **Overall**  N = 312^1^ | **RCT**  N = 103^1^ | **MA**  N = 98^1^ | **Other**  N = 111^1^ |
| --- | --- | --- | --- | --- |
| Journal |  |  |  |  |
| Ann Intern Med | 30 (9.6%) | 11 (11%) | 10 (10%) | 9 (8.1%) |
| BMC Med | 36 (12%) | 10 (9.7%) | 16 (16%) | 10 (9.0%) |
| BMJ | 31 (9.9%) | 10 (9.7%) | 13 (13%) | 8 (7.2%) |
| CMAJ | 17 (5.4%) | 3 (2.9%) | 4 (4.1%) | 10 (9.0%) |
| JAMA | 32 (10%) | 11 (11%) | 11 (11%) | 10 (9.0%) |
| JAMA Netw Open | 43 (14%) | 13 (13%) | 13 (13%) | 17 (15%) |
| Lancet | 34 (11%) | 12 (12%) | 10 (10%) | 12 (11%) |
| N Engl J Med | 23 (7.4%) | 14 (14%) | 0 (0%) | 9 (8.1%) |
| Nat Med | 32 (10%) | 10 (9.7%) | 7 (7.1%) | 15 (14%) |
| PLoS Med | 34 (11%) | 9 (8.7%) | 14 (14%) | 11 (9.9%) |
| Registration | 189 (61%) | 101 (98%) | 68 (69%) | 20 (18%) |
| Data Sharing |  |  |  |  |
| no sharing | 70 (22%) | 17 (17%) | 23 (23%) | 30 (27%) |
| open access | 56 (18%) | 6 (5.8%) | 35 (36%) | 15 (14%) |
| under request | 186 (60%) | 80 (78%) | 40 (41%) | 66 (59%) |
| Open Access Sharing | 257 (82%) | 82 (80%) | 83 (85%) | 92 (83%) |
| Code Sharing |  |  |  |  |
| no sharing | 237 (76%) | 86 (83%) | 70 (71%) | 81 (73%) |
| open access | 40 (13%) | 3 (2.9%) | 15 (15%) | 22 (20%) |
| under request | 35 (11%) | 14 (14%) | 13 (13%) | 8 (7.2%) |
| Publication 1year RCT | 38 (35%) | 37 (37%) | 0 (NA%) | 1 (11%) |
| Protocol |  |  |  |  |
| no sharing | 129 (41%) | 11 (11%) | 32 (33%) | 86 (77%) |
| open access | 172 (55%) | 87 (84%) | 63 (64%) | 22 (20%) |
| under request | 11 (3.5%) | 5 (4.9%) | 3 (3.1%) | 3 (2.7%) |
| Statistical Analysis Plan |  |  |  |  |
| no sharing | 247 (79%) | 51 (50%) | 94 (96%) | 102 (92%) |
| open access | 60 (19%) | 48 (47%) | 4 (4.1%) | 8 (7.2%) |
| under request | 5 (1.6%) | 4 (3.9%) | 0 (0%) | 1 (0.9%) |
| Reporting guideline | 148 (47%) | 41 (40%) | 68 (69%) | 39 (35%) |
| Preprint | 1 (0.3%) | 1 (1.0%) | 0 (0%) | 0 (0%) |
| Author contribution | 289 (93%) | 88 (85%) | 96 (98%) | 105 (95%) |
| COI | 312 (100%) | 103 (100%) | 98 (100%) | 111 (100%) |
| ORCID | 137 (44%) | 46 (45%) | 44 (45%) | 47 (42%) |
| Funding statement | 306 (98%) | 103 (100%) | 95 (97%) | 108 (97%) |
| ^1^n (%) | | | | |

*Abbreviation:*

*RCT: Randomized Control Trial,*

*MA: Meta-analysis,*

*COI: Conflict of interest*

*ORCID: Open Researcher and Contributor ID*

*n (%) : number (percentage)*

**Web Appendix 10: Description of the accuracy of the automatic tools (Sensitivity, Specificity, and F1).**

*Abbreviation: CI: Confidence intervale, RCT: Randomised control trial,*

| Tool | Metric | RCT | Registration | Intention to share data | Open data sharing | Open access publishing | Code sharing | Publication 1year (for RCT) | Protocol sharing | Statistical analysis plan sharing | Utilization of reporting guideline | Preprint | Author contribution | COI statement | Reporting ORCID identifiers | Funding statement |
| --- | --- | --- | --- | --- | --- | --- | --- | --- | --- | --- | --- | --- | --- | --- | --- | --- |
| CTregistries | Sensibility (CI 95%) |  | 0.57 (0.49 - 0.64) |  |  |  |  |  |  |  |  |  |  |  |  |  |
|  | Specificity (CI 95%) |  | 0.84 (0.76 - 0.90) |  |  |  |  |  |  |  |  |  |  |  |  |  |
|  | F1-score |  | 0.68 |  |  |  |  |  |  |  |  |  |  |  |  |  |
| ContriBOT | Sensibility (CI 95%) |  |  |  |  |  |  |  |  |  |  |  | 0.87 (0.82 - 0.90) |  | 0.86 (0.79 - 0.91) |  |
|  | Specificity (CI 95%) |  |  |  |  |  |  |  |  |  |  |  | 0.78 (0.56 - 0.93) |  | 0.99 (0.96 - 1.00) |  |
|  | F1-score |  |  |  |  |  |  |  |  |  |  |  | 0.92 |  | 0.92 |  |
| Data Seer | Sensibility (CI 95%) |  | 0.80 (0.72 - 0.85) | 0.40 (0.33 - 0.47) |  |  | 0.19 (0.10 - 0.31) |  |  |  |  | 0.00 (0.00 - 0.97) |  |  |  |  |
|  | Specificity (CI 95%) |  | 0.93 (0.87 - 0.97) | 0.86 (0.75 - 0.93) |  |  | 0.99 (0.96 - 1.00) |  |  |  |  | 1.00 (0.99 - 1.00) |  |  |  |  |
|  | F1-score |  | 0.86 | 0.55 |  |  | 0.31 |  |  |  |  | incalculable |  |  |  |  |
| Llama 3.3 70B | Sensibility (CI 95%) | 0.93 (0.86 - 0.97) | 0.72 (0.65 - 0.78) | 0.83 (0.78 - 0.88) | 0.50 (0.36 - 0.64) | 0.41 (0.35 - 0.47) | 0.71 (0.59 - 0.81) | 0.46 (0.30 - 0.63) | 0.79 (0.73 - 0.85) | 0.95 (0.87 - 0.99) | 0.74 (0.67 - 0.81) | 1.00 (0.03 - 1.00) | 0.82 (0.77 - 0.86) | 0.78 (0.73 - 0.82) | 0.03 (0.01 - 0.07) | 0.86 (0.82 - 0.90) |
|  | Specificity (CI 95%) | 0.89 (0.84 - 0.93) | 0.92 (0.86 - 0.96) | 0.53 (0.41 - 0.65) | 0.93 (0.89 - 0.96) | 0.87 (0.76 - 0.95) | 0.71 (0.65 - 0.77) | 0.64 (0.52 - 0.75) | 0.45 (0.36 - 0.54) | 0.35 (0.29 - 0.42) | 0.82 (0.76 - 0.88) | 0.96 (0.93 - 0.98) | 0.04 (0.00 - 0.22) | NaN (0.00 - 1.00) | 1.00 (0.98 - 1.00) | 0.67 (0.22 - 0.96) |
|  | F1-score | 0.87 | 0.81 | 0.85 | 0.55 | 0.57 | 0.54 | 0.43 | 0.73 | 0.43 | 0.77 | 0.14 | 0.87 | 0.88 | 0.06 | 0.92 |
| ODDPub | Sensibility (CI 95%) |  |  | 0.70 (0.64 - 0.76) | 0.27 (0.16 - 0.40) |  | 0.45 (0.34 - 0.57) |  |  |  |  |  |  |  |  |  |
|  | Specificity (CI 95%) |  |  | 0.61 (0.49 - 0.73) | 0.97 (0.94 - 0.99) |  | 0.89 (0.84 - 0.92) |  |  |  |  |  |  |  |  |  |
|  | F1-score |  |  | 0.77 | 0.38 |  | 0.50 |  |  |  |  |  |  |  |  |  |
| SciScore | Sensibility (CI 95%) | 0.91 (0.82 - 0.96) | 0.09 (0.05 - 0.15) | 0.27 (0.21 - 0.34) |  |  | 0.11 (0.04 - 0.21) |  |  |  |  |  |  |  |  |  |
|  | Specificity (CI 95%) | 0.29 (0.22 - 0.36) | 0.99 (0.94 - 1.00) | 0.78 (0.64 - 0.88) |  |  | 0.99 (0.97 - 1.00) |  |  |  |  |  |  |  |  |  |
|  | F1-score | 0.53 | 0.17 | 0.41 |  |  | 0.19 |  |  |  |  |  |  |  |  |  |
| TRNscreener | Sensibility (CI 95%) |  | 0.57 (0.50 - 0.64) |  |  |  |  |  |  |  |  |  |  |  |  |  |
|  | Specificity (CI 95%) |  | 0.84 (0.76 - 0.90) |  |  |  |  |  |  |  |  |  |  |  |  |  |
|  | F1-score |  | 0.68 |  |  |  |  |  |  |  |  |  |  |  |  |  |
| rTransparent | Sensibility (CI 95%) |  | 0.77 (0.70 - 0.83) | 0.74 (0.68 - 0.80) | 0.25 (0.14 - 0.38) |  | 0.39 (0.28 - 0.51) |  |  |  |  |  |  | 1.00 (0.99 - 1.00) |  | 0.99 (0.97 - 1.00) |
|  | Specificity (CI 95%) |  | 0.93 (0.88 - 0.97) | 0.59 (0.46 - 0.70) | 0.96 (0.92 - 0.98) |  | 0.96 (0.92 - 0.98) |  |  |  |  |  |  | incalculable |  | 0.67 (0.22 - 0.96) |
|  | F1-score |  | 0.85 | 0.80 | 0.35 |  | 0.51 |  |  |  |  |  |  | 1 |  | 0.99 |
| Unpaywall | Sensibility (CI 95%) |  |  |  |  | 0.99 (0.96-1.0) |  |  |  |  |  |  |  |  |  |  |
|  | Specificity (CI 95%) |  |  |  |  | 0.36 (0.11-0.69) |  |  |  |  |  |  |  |  |  |  |
|  | F1-score |  |  |  |  | 0.98 |  |  |  |  |  |  |  |  |  |  |

**Web Appendix 11-24: Automatic tool performance depending on the study type.**

### ***Sensitivity analysis of the performance of automated tools by study type: MA***

| Tool | Metric | RCT | Registration | Intention to share data | Open data sharing | Open access publishing | Code sharing | Protocol sharing | Statistical analysis plan sharing | Utilization of reporting guideline | Preprint | Author contribution | COI statement | Reporting ORCID identifiers | Funding statement |
| --- | --- | --- | --- | --- | --- | --- | --- | --- | --- | --- | --- | --- | --- | --- | --- |
| CTregistries | Sensibility (CI 95%) |  | 0.28 (0.18 - 0.40) |  |  |  |  |  |  |  |  |  |  |  |  |
|  | Specificity (CI 95%) |  | 0.73 (0.54 - 0.88) |  |  |  |  |  |  |  |  |  |  |  |  |
|  | F1-score |  | 0.400 |  |  |  |  |  |  |  |  |  |  |  |  |
| ContriBOT | Sensibility (CI 95%) |  |  |  |  |  |  |  |  |  |  | 0.88 (0.79 - 0.93) |  | 0.95 (0.85 - 0.99) |  |
|  | Specificity (CI 95%) |  |  |  |  |  |  |  |  |  |  | 0.00 (0.00 - 0.84) |  | 0.98 (0.90 - 1.00) |  |
|  | F1-score |  |  |  |  |  |  |  |  |  |  | 0.923 |  | 0.966 |  |
| Data Seer | Sensibility (CI 95%) |  | 0.80 (0.68 - 0.90) | 0.26 (0.16 - 0.38) |  |  | 0.21 (0.07 - 0.42) |  |  |  | incalculable |  |  |  |  |
|  | Specificity (CI 95%) |  | 0.96 (0.82 - 1.00) | 0.95 (0.77 - 1.00) |  |  | 1.00 (0.94 - 1.00) |  |  |  | 1.00 (0.96 - 1.00) |  |  |  |  |
|  | F1-score |  | 0.882 | 0.405 |  |  | 0.345 |  |  |  | incalculable |  |  |  |  |
| Llama 3.3 70B | Sensibility (CI 95%) | incalculable | 0.72 (0.60 - 0.82) | 0.73 (0.62 - 0.83) | 0.46 (0.29 - 0.63) | 0.41 (0.30 - 0.52) | 0.64 (0.44 - 0.81) | 0.68 (0.56 - 0.79) | 1.00 (0.40 - 1.00) | 0.84 (0.73 - 0.92) | incalculable | 0.88 (0.79 - 0.93) | 0.67 (0.57 - 0.76) | 0.07 (0.01 - 0.19) | 0.76 (0.66 - 0.84) |
|  | Specificity (CI 95%) | 0.94 (0.87 - 0.98) | 0.87 (0.69 - 0.96) | 0.74 (0.52 - 0.90) | 0.94 (0.85 - 0.98) | 0.87 (0.60 - 0.98) | 0.83 (0.72 - 0.91) | 0.59 (0.41 - 0.76) | 0.40 (0.30 - 0.51) | 0.80 (0.61 - 0.92) | 0.92 (0.85 - 0.96) | 0.00 (0.00 - 0.84) | incalculable | 1.00 (0.93 - 1.00) | 0.67 (0.09 - 0.99) |
|  | F1-score | incalculable | 0.810 | 0.809 | 0.58 | 0.571 | 0.621 | 0.726 | 0.125 | 0.870 | incalculable | 0.923 | 0.805 | 0.128 | 0.857 |
| ODDPub | Sensibility (CI 95%) |  |  | 0.65 (0.53 - 0.76) | 0.26 (0.12 - 0.43) |  | 0.46 (0.28 - 0.66) |  |  |  |  |  |  |  |  |
|  | Specificity (CI 95%) |  |  | 0.78 (0.56 - 0.93) | 0.97 (0.89 - 1.00) |  | 0.96 (0.88 - 0.99) |  |  |  |  |  |  |  |  |
|  | F1-score |  |  | 0.760 | 0.38 |  | 0.591 |  |  |  |  |  |  |  |  |
| SciScore | Sensibility (CI 95%) |  | 0.00 (0.00 - 0.06) | 0.40 (0.28 - 0.53) |  |  | 0.12 (0.03 - 0.31) |  |  |  |  |  |  |  |  |
|  | Specificity (CI 95%) |  | 1.00 (0.87 - 1.00) | 0.68 (0.43 - 0.87) |  |  | 0.98 (0.91 - 1.00) |  |  |  |  |  |  |  |  |
|  | F1-score |  | incalculable | 0.536 |  |  | 0.207 |  |  |  |  |  |  |  |  |
| TNRscreener | Sensibility (CI 95%) |  | 0.29 (0.19 - 0.42) |  |  |  |  |  |  |  |  |  |  |  |  |
|  | Specificity (CI 95%) |  | 0.73 (0.54 - 0.88) |  |  |  |  |  |  |  |  |  |  |  |  |
|  | F1-score |  | 0.417 |  |  |  |  |  |  |  |  |  |  |  |  |
| rTransparent | Sensibility (CI 95%) |  | 0.81 (0.70 - 0.89) | 0.69 (0.58 - 0.79) | 0.23 (0.1 - 0.4) |  | 0.36 (0.19 - 0.56) |  |  |  |  |  | 1.00 (0.96 - 1.00) |  | 0.96 (0.90 - 0.99) |
|  | Specificity (CI 95%) |  | 0.90 (0.73 - 0.98) | 0.78 (0.56 - 0.93) | 0.98 (0.91 - 1) |  | 0.94 (0.86 - 0.98) |  |  |  |  |  | incalculable |  | 0.33 (0.01 - 0.91) |
|  | F1-score |  | 0.873 | 0.788 | 0.36 |  | 0.476 |  |  |  |  |  | 1.000 |  | 0.968 |
| Unpaywall | Sensibility (CI 95%) |  |  |  |  | 0.96 (0.88-0.99) |  |  |  |  |  |  |  |  |  |
|  | Specificity (CI 95%) |  |  |  |  | 0.67 (0.09-0.99) |  |  |  |  |  |  |  |  |  |
|  | F1-score |  |  |  |  | 0.97 |  |  |  |  |  |  |  |  |  |

### ***Sensitivity analysis of the performance of automated tools by study type: RA***

| Tool | Metric | RCT | Registration | Intention to share data | Open data sharing | Open access publishing | Code sharing | Protocol sharing | Statistical analysis plan sharing | Utilization of reporting guideline | Preprint | Author contribution | COI statement | Reporting ORCID identifiers | Funding statement |
| --- | --- | --- | --- | --- | --- | --- | --- | --- | --- | --- | --- | --- | --- | --- | --- |
| CTregistries | Sensibility (CI 95%) |  | 0.70 (0.46 - 0.88) |  |  |  |  |  |  |  |  |  |  |  |  |
|  | Specificity (CI 95%) |  | 0.87 (0.78 - 0.93) |  |  |  |  |  |  |  |  |  |  |  |  |
|  | F1-score |  | 0.609 |  |  |  |  |  |  |  |  |  |  |  |  |
| ContriBOT | Sensibility (CI 95%) |  |  |  |  |  |  |  |  |  |  | 0.87 (0.79 - 0.93) |  | 0.85 (0.72 - 0.94) |  |
|  | Specificity (CI 95%) |  |  |  |  |  |  |  |  |  |  | 1.00 (0.54 - 1.00) |  | 0.98 (0.92 - 1.00) |  |
|  | F1-score |  |  |  |  |  |  |  |  |  |  | 0.929 |  | 0.909 |  |
| Data Seer | Sensibility (CI 95%) |  | 0.71 (0.44 - 0.90) | 0.58 (0.46 - 0.71) |  |  | 0.25 (0.10 - 0.47) |  |  |  | incalculable |  |  |  |  |
|  | Specificity (CI 95%) |  | 0.92 (0.83 - 0.97) | 0.81 (0.61 - 0.93) |  |  | 0.99 (0.92 - 1.00) |  |  |  | 1.00 (0.96 - 1.00) |  |  |  |  |
|  | F1-score |  | 0.686 | 0.704 |  |  | 0.387 |  |  |  | incalculable |  |  |  |  |
| Llama 3.3 70B | Sensibility (CI 95%) | incalculable | 0.70 (0.46 - 0.88) | 0.86 (0.77 - 0.93) | 0.47 (0.21 - 0.73) | 0.40 (0.30 - 0.51) | 0.80 (0.61 - 0.92) | 0.88 (0.69 - 0.97) | 1.00 (0.66 - 1.00) | 0.67 (0.50 - 0.81) | incalculable | 0.77 (0.68 - 0.85) | 0.79 (0.71 - 0.86) | 0.00 (0.00 - 0.08) | 0.93 (0.86 - 0.97) |
|  | Specificity (CI 95%) | 0.86 (0.78 - 0.92) | 0.93 (0.86 - 0.98) | 0.57 (0.37 - 0.75) | 0.9 (0.82 - 0.95) | 0.79 (0.54 - 0.94) | 0.70 (0.59 - 0.80) | 0.43 (0.32 - 0.54) | 0.38 (0.29 - 0.48) | 0.88 (0.78 - 0.94) | 0.96 (0.91 - 0.99) | 0.17 (0.00 - 0.64) | incalculable | 1.00 (0.94 - 1.00) | 0.67 (0.09 - 0.99) |
|  | F1-score | incalculable | 0.700 | 0.854 | 0.44 | 0.556 | 0.615 | 0.458 | 0.222 | 0.703 | incalculable | 0.848 | 0.884 | incalculable | 0.957 |
| ODDPub | Sensibility (CI 95%) |  |  | 0.67 (0.55 - 0.77) | 0.2 (0.04 - 0.48) |  | 0.60 (0.41 - 0.77) |  |  |  |  |  |  |  |  |
|  | Specificity (CI 95%) |  |  | 0.63 (0.44 - 0.80) | 0.94 (0.87 - 0.98) |  | 0.89 (0.80 - 0.95) |  |  |  |  |  |  |  |  |
|  | F1-score |  |  | 0.740 | 0.25 |  | 0.632 |  |  |  |  |  |  |  |  |
| SciScore | Sensibility (CI 95%) |  | 0.19 (0.04 - 0.46) | 0.22 (0.12 - 0.34) |  |  | 0.12 (0.02 - 0.30) |  |  |  |  |  |  |  |  |
|  | Specificity (CI 95%) |  | 0.98 (0.92 - 1.00) | 0.80 (0.56 - 0.94) |  |  | 1.00 (0.93 - 1.00) |  |  |  |  |  |  |  |  |
|  | F1-score |  | 0.300 | 0.338 |  |  | 0.207 |  |  |  |  |  |  |  |  |
| TNRscreener | Sensibility (CI 95%) |  | 0.70 (0.46 - 0.88) |  |  |  |  |  |  |  |  |  |  |  |  |
|  | Specificity (CI 95%) |  | 0.87 (0.78 - 0.93) |  |  |  |  |  |  |  |  |  |  |  |  |
|  | F1-score |  | 0.609 |  |  |  |  |  |  |  |  |  |  |  |  |
| rTransparent | Sensibility (CI 95%) |  | 0.70 (0.46 - 0.88) | 0.73 (0.62 - 0.82) | 0.2 (0.04 - 0.48) |  | 0.60 (0.41 - 0.77) |  |  |  |  |  | 1.00 (0.97 - 1.00) |  | 1.00 (0.97 - 1.00) |
|  | Specificity (CI 95%) |  | 0.95 (0.88 - 0.98) | 0.57 (0.37 - 0.75) | 0.9 (0.82 - 0.95) |  | 0.94 (0.86 - 0.98) |  |  |  |  |  | incalculable |  | 1.00 (0.29 - 1.00) |
|  | F1-score |  | 0.718 | 0.771 | 0.21 |  | 0.679 |  |  |  |  |  | 1.000 |  | 1.000 |
| Unpaywall | Sensibility (CI 95%) |  |  |  |  | 1 (0.95-1) |  |  |  |  |  |  |  |  |  |
|  | Specificity (CI 95%) |  |  |  |  | 0.29 (0.04-0.70) |  |  |  |  |  |  |  |  |  |
|  | F1-score |  |  |  |  | 0.97 |  |  |  |  |  |  |  |  |  |

### ***Sensitivity analysis of the performance of automated tools by study type: RCT***

| Tool | Metric | RCT | Registration | Intention to share data | Open data sharing | Open access publishing | Code sharing | Publication 1year (for RCT) | Protocol sharing | Statistical analysis plan sharing | Utilization of reporting guideline | Preprint | Author contribution | COI statement | Reporting ORCID identifiers | Funding statement |
| --- | --- | --- | --- | --- | --- | --- | --- | --- | --- | --- | --- | --- | --- | --- | --- | --- |
| CTregistries | Sensibility (CI 95%) |  | 0.73 (0.64 - 0.82) |  |  |  |  |  |  |  |  |  |  |  |  |  |
|  | Specificity (CI 95%) |  | 1.00 (0.16 - 1.00) |  |  |  |  |  |  |  |  |  |  |  |  |  |
|  | F1-score |  | 0.846 |  |  |  |  |  |  |  |  |  |  |  |  |  |
| ContriBOT | Sensibility (CI 95%) |  |  |  |  |  |  |  |  |  |  |  | 0.85 (0.76 - 0.92) |  | 0.78 (0.64 - 0.89) |  |
|  | Specificity (CI 95%) |  |  |  |  |  |  |  |  |  |  |  | 0.80 (0.52 - 0.96) |  | 1.00 (0.94 - 1.00) |  |
|  | F1-score |  |  |  |  |  |  |  |  |  |  |  | 0.904 |  | 0.878 |  |
| Data Seer | Sensibility (CI 95%) |  | 0.81 (0.71 - 0.88) | 0.35 (0.24 - 0.47) |  |  | 0.07 (0.00 - 0.32) |  |  |  |  | 0.00 (0.00 - 0.97) |  |  |  |  |
|  | Specificity (CI 95%) |  | 1.00 (0.16 - 1.00) | 0.81 (0.54 - 0.96) |  |  | 0.99 (0.93 - 1.00) |  |  |  |  | 1.00 (0.96 - 1.00) |  |  |  |  |
|  | F1-score |  | 0.893 | 0.505 |  |  | 0.118 |  |  |  |  | incalculable |  |  |  |  |
| Llama 3..3 70B | Sensibility (CI 95%) | 0.93 (0.86 - 0.97) | 0.72 (0.62 - 0.81) | 0.88 (0.80 - 0.94) | 0.83 (0.36 - 1) | 0.41 (0.31 - 0.53) | 0.65 (0.38 - 0.86) | 0.45 (0.29 - 0.62) | 0.85 (0.76 - 0.91) | 0.94 (0.84 - 0.99) | 0.66 (0.49 - 0.80) | 1.00 (0.03 - 1.00) | 0.82 (0.72 - 0.89) | 0.86 (0.78 - 0.92) | 0.02 (0.00 - 0.12) | 0.89 (0.82 - 0.95) |
|  | Specificity (CI 95%) | incalculable | 1.00 (0.16 - 1.00) | 0.18 (0.04 - 0.43) | 0.96 (0.9 - 0.99) | 0.95 (0.76 - 1.00) | 0.62 (0.51 - 0.72) | 0.63 (0.50 - 0.75) | 0.18 (0.02 - 0.52) | 0.20 (0.10 - 0.33) | 0.77 (0.65 - 0.87) | 1.00 (0.96 - 1.00) | 0.00 (0.00 - 0.22) | incalculable | 1.00 (0.94 - 1.00) | incalculable |
|  | F1-score | 0.965 | 0.839 | 0.864 | 0.67 | 0.581 | 0.361 | 0.430 | 0.872 | 0.690 | 0.659 | 1.000 | 0.823 | 0.927 | 0.043 | 0.944 |
| ODDPub | Sensibility (CI 95%) |  |  | 0.77 (0.66 - 0.85) | 0.50 (0.12 - 0.88) |  | 0.18 (0.04 - 0.43) |  |  |  |  |  |  |  |  |  |
|  | Specificity (CI 95%) |  |  | 0.35 (0.14 - 0.62) | 1.00 (0.96 - 1.00) |  | 0.83 (0.73 - 0.90) |  |  |  |  |  |  |  |  |  |
|  | F1-score |  |  | 0.810 | 0.67 |  | 0.171 |  |  |  |  |  |  |  |  |  |
| SciScore | Sensibility (CI 95%) |  | 0.15 (0.08 - 0.25) | 0.20 (0.11 - 0.31) |  |  | 0.07 (0.00 - 0.32) |  |  |  |  |  |  |  |  |  |
|  | Specificity (CI 95%) |  | 1.00 (0.16 - 1.00) | 0.91 (0.59 - 1.00) |  |  | 1.00 (0.94 - 1.00) |  |  |  |  |  |  |  |  |  |
|  | F1-score |  | 0.256 | 0.325 |  |  | 0.125 |  |  |  |  |  |  |  |  |  |
| TNRscreener | Sensibility (CI 95%) |  | 0.73 (0.64 - 0.82) |  |  |  |  |  |  |  |  |  |  |  |  |  |
|  | Specificity (CI 95%) |  | 1.00 (0.16 - 1.00) |  |  |  |  |  |  |  |  |  |  |  |  |  |
|  | F1-score |  | 0.846 |  |  |  |  |  |  |  |  |  |  |  |  |  |
| rTransparent | Sensibility (CI 95%) |  | 0.75 (0.66 - 0.83) | 0.80 (0.70 - 0.88) | 0.5 (0.12 - 0.88) |  | 0.06 (0.00 - 0.29) |  |  |  |  |  |  | 1.00 (0.96 - 1.00) |  | 1.00 (0.96 - 1.00) |
|  | Specificity (CI 95%) |  | 1.00 (0.16 - 1.00) | 0.35 (0.14 - 0.62) | 1 (0.96 - 1) |  | 0.99 (0.94 - 1.00) |  |  |  |  |  |  | incalculable |  | incalculable |
|  | F1-score |  | 0.859 | 0.831 | 0.67 |  | 0.105 |  |  |  |  |  |  | 1.000 |  | 1.000 |
| Unpaywall | Sensibility (CI 95%) |  |  |  |  | 1 (0.94-1) |  |  |  |  |  |  |  |  |  |  |
|  | Specificity (CI 95%) |  |  |  |  | 0.00 (0.00-0.98) |  |  |  |  |  |  |  |  |  |  |
|  | F1-score |  |  |  |  | 0.99 |  |  |  |  |  |  |  |  |  |  |

### ***Sensitivity analysis of the performance of automated tools by journal: Annals of internal Medicine***

| Tool | Metric | RCT | Registration | Intention to share data | Open data sharing | Open access publishing | Code sharing | Publication 1year (for RCT) | Protocol sharing | Statistical analysis plan sharing | Utilization of reporting guideline | Preprint | Author contribution | COI statement | Reporting ORCID identifiers | Funding statement |
| --- | --- | --- | --- | --- | --- | --- | --- | --- | --- | --- | --- | --- | --- | --- | --- | --- |
| CTregistries | Sensibility (CI 95%) |  | 0.61 (0.36 - 0.83) |  |  |  |  |  |  |  |  |  |  |  |  |  |
|  | Specificity (CI 95%) |  | 0.83 (0.52 - 0.98) |  |  |  |  |  |  |  |  |  |  |  |  |  |
|  | F1-score |  | 0.710 |  |  |  |  |  |  |  |  |  |  |  |  |  |
| ContriBOT | Sensibility (CI 95%) |  |  |  |  |  |  |  |  |  |  |  | 1.00 (0.88 - 1.00) |  | incalculable |  |
|  | Specificity (CI 95%) |  |  |  |  |  |  |  |  |  |  |  | 0.00 (0.00 - 0.84) |  | 1.00 (0.88 - 1.00) |  |
|  | F1-score |  |  |  |  |  |  |  |  |  |  |  | 0.966 |  | incalculable |  |
| Data Seer | Sensibility (CI 95%) |  | 0.78 (0.52 - 0.94) | 0.10 (0.01 - 0.32) |  |  | 0.00 (0.00 - 0.20) |  |  |  |  | 0.00 (0.00 - 0.97) |  |  |  |  |
|  | Specificity (CI 95%) |  | 1.00 (0.72 - 1.00) | 1.00 (0.66 - 1.00) |  |  | 1.00 (0.74 - 1.00) |  |  |  |  | 1.00 (0.88 - 1.00) |  |  |  |  |
|  | F1-score |  | 0.875 | 0.182 |  |  | incalculable |  |  |  |  | incalculable |  |  |  |  |
| Llama 3.3 70B | Sensibility (CI 95%) | 1.00 (0.72 - 1.00) | 0.78 (0.52 - 0.94) | 0.57 (0.34 - 0.78) | 0.5 (0.07 - 0.93) | 0.00 (0.00 - 0.71) | 0.56 (0.31 - 0.78) | 0.50 (0.07 - 0.93) | 0.87 (0.66 - 0.97) | 0.83 (0.36 - 1.00) | 0.75 (0.35 - 0.97) | 1.00 (0.03 - 1.00) | 0.79 (0.59 - 0.92) | 0.83 (0.65 - 0.94) | incalculable | 0.87 (0.69 - 0.96) |
|  | Specificity (CI 95%) | 0.95 (0.74 - 1.00) | 0.92 (0.62 - 1.00) | 0.78 (0.40 - 0.97) | 1 (0.87 - 1) | 0.93 (0.76 - 0.99) | 0.75 (0.43 - 0.95) | 0.88 (0.47 - 1.00) | 0.71 (0.29 - 0.96) | 0.21 (0.07 - 0.42) | 0.86 (0.65 - 0.97) | 0.97 (0.82 - 1.00) | 0.00 (0.00 - 0.84) | incalculable | 1.00 (0.88 - 1.00) | incalculable |
|  | F1-score | 0.957 | 0.848 | 0.686 | 0.67 | incalculable | 0.645 | 0.571 | 0.889 | 0.333 | 0.706 | 0.667 | 0.846 | 0.909 | incalculable | 0.929 |
| ODDPub | Sensibility (CI 95%) |  |  | 0.48 (0.26 - 0.70) | 0.5 (0.07 - 0.93) |  | 0.33 (0.13 - 0.59) |  |  |  |  |  |  |  |  |  |
|  | Specificity (CI 95%) |  |  | 0.67 (0.30 - 0.93) | 1 (0.87 - 1) |  | 0.92 (0.62 - 1.00) |  |  |  |  |  |  |  |  |  |
|  | F1-score |  |  | 0.588 | 0.67 |  | 0.480 |  |  |  |  |  |  |  |  |  |
| SciScore | Sensibility (CI 95%) |  | 0.12 (0.01 - 0.36) | 0.19 (0.05 - 0.42) |  |  | 0.00 (0.00 - 0.19) |  |  |  |  |  |  |  |  |  |
|  | Specificity (CI 95%) |  | 1.00 (0.74 - 1.00) | 0.75 (0.35 - 0.97) |  |  | 1.00 (0.72 - 1.00) |  |  |  |  |  |  |  |  |  |
|  | F1-score |  | 0.211 | 0.296 |  |  |  |  |  |  |  |  |  |  |  |  |
| TNRscreener | Sensibility (CI 95%) |  | 0.61 (0.36 - 0.83) |  |  |  |  |  |  |  |  |  |  |  |  |  |
|  | Specificity (CI 95%) |  | 0.83 (0.52 - 0.98) |  |  |  |  |  |  |  |  |  |  |  |  |  |
|  | F1-score |  | 0.710 |  |  |  |  |  |  |  |  |  |  |  |  |  |
| rTransparent | Sensibility (CI 95%) |  | 0.61 (0.36 - 0.83) | 0.57 (0.34 - 0.78) | 0.5 (0.07 - 0.93) |  | 0.22 (0.06 - 0.48) |  |  |  |  |  |  | 1.00 (0.88 - 1.00) |  | 1.00 (0.88 - 1.00) |
|  | Specificity (CI 95%) |  | 1.00 (0.74 - 1.00) | 0.67 (0.30 - 0.93) | 1 (0.87 - 1) |  | 1.00 (0.74 - 1.00) |  |  |  |  |  |  | incalculable |  | incalculable |
|  | F1-score |  | 0.759 | 0.667 | 0.67 |  | 0.364 |  |  |  |  |  |  | 1.000 |  | 1.000 |
| Unpaywall | Sensibility (CI 95%) |  |  |  |  | incalculable |  |  |  |  |  |  |  |  |  |  |
|  | Specificity (CI 95%) |  |  |  |  | incalculable |  |  |  |  |  |  |  |  |  |  |
|  | F1-score |  |  |  |  | incalculable |  |  |  |  |  |  |  |  |  |  |

### ***Sensitivity analysis of the performance of automated tools by journal: BMC Medicine***

| Tool | Metric | RCT | Registration | Intention to share data | Open data sharing | Open access publishing | Code sharing | Publication 1year (for RCT) | Protocol sharing | Statistical analysis plan sharing | Utilization of reporting guideline | Preprint | Author contribution | COI statement | Reporting ORCID identifiers | Funding statement |
| --- | --- | --- | --- | --- | --- | --- | --- | --- | --- | --- | --- | --- | --- | --- | --- | --- |
| CTregistries | Sensibility (CI 95%) |  | 0.33 (0.16 - 0.55) |  |  |  |  |  |  |  |  |  |  |  |  |  |
|  | Specificity (CI 95%) |  | 0.92 (0.62 - 1.00) |  |  |  |  |  |  |  |  |  |  |  |  |  |
|  | F1-score |  | 0.485 |  |  |  |  |  |  |  |  |  |  |  |  |  |
| ContriBOT | Sensibility (CI 95%) |  |  |  |  |  |  |  |  |  |  |  | 1.00 (0.90 - 1.00) |  | 0.96 (0.80 - 1.00) |  |
|  | Specificity (CI 95%) |  |  |  |  |  |  |  |  |  |  |  | incalculable |  | 1.00 (0.72 - 1.00) |  |
|  | F1-score |  |  |  |  |  |  |  |  |  |  |  | 1.000 |  | 0.980 |  |
| Data Seer | Sensibility (CI 95%) |  | 0.88 (0.68 - 0.97) | 0.42 (0.25 - 0.61) |  |  | 0.33 (0.04 - 0.78) |  |  |  |  | incalculable |  |  |  |  |
|  | Specificity (CI 95%) |  | 1.00 (0.74 - 1.00) | 0.67 (0.09 - 0.99) |  |  | 1.00 (0.88 - 1.00) |  |  |  |  | 1.00 (0.90 - 1.00) |  |  |  |  |
|  | F1-score |  | 0.933 | 0.583 |  |  | 0.500 |  |  |  |  | incalculable |  |  |  |  |
| Llama 3.3 70B | Sensibility (CI 95%) | 1.00 (0.69 - 1.00) | 0.54 (0.33 - 0.74) | 0.91 (0.76 - 0.98) | 0.67 (0.3 - 0.93) | 0.81 (0.64 - 0.92) | 0.33 (0.04 - 0.78) | 0.00 (0.00 - 0.84) | 0.61 (0.36 - 0.83) | 1.00 (0.16 - 1.00) | 0.71 (0.44 - 0.90) | incalculable | 0.83 (0.67 - 0.94) | 1.00 (0.90 - 1.00) | 0.00 (0.00 - 0.14) | 0.89 (0.74 - 0.97) |
|  | Specificity (CI 95%) | 0.88 (0.70 - 0.98) | 1.00 (0.74 - 1.00) | 0.67 (0.09 - 0.99) | 0.93 (0.76 - 0.99) | incalculable | 0.83 (0.65 - 0.94) | 1.00 (0.66 - 1.00) | 0.56 (0.31 - 0.78) | 0.41 (0.25 - 0.59) | 0.95 (0.74 - 1.00) | 0.92 (0.78 - 0.98) | incalculable | incalculable | 1.00 (0.72 - 1.00) | incalculable |
|  | F1-score | 0.870 | 0.703 | 0.938 | 0.71 | 0.892 | 0.308 | incalculable | 0.595 | 0.167 | 0.800 | incalculable | 0.909 | 1.000 | incalculable | 0.941 |
| ODDPub | Sensibility (CI 95%) |  |  | 0.82 (0.65 - 0.93) | 0.11 (0 - 0.48) |  | 0.67 (0.22 - 0.96) |  |  |  |  |  |  |  |  |  |
|  | Specificity (CI 95%) |  |  | 0.67 (0.09 - 0.99) | 1 (0.87 - 1) |  | 1.00 (0.88 - 1.00) |  |  |  |  |  |  |  |  |  |
|  | F1-score |  |  | 0.885 | 0.20 |  | 0.800 |  |  |  |  |  |  |  |  |  |
| SciScore | Sensibility (CI 95%) |  | 0.05 (0.00 - 0.24) | 0.10 (0.02 - 0.27) |  |  | 0.00 (0.00 - 0.46) |  |  |  |  |  |  |  |  |  |
|  | Specificity (CI 95%) |  | 1.00 (0.74 - 1.00) | 1.00 (0.29 - 1.00) |  |  | 1.00 (0.87 - 1.00) |  |  |  |  |  |  |  |  |  |
|  | F1-score |  | 0.091 | 0.182 |  |  |  |  |  |  |  |  |  |  |  |  |
| TNRscreener | Sensibility (CI 95%) |  | 0.33 (0.16 - 0.55) |  |  |  |  |  |  |  |  |  |  |  |  |  |
|  | Specificity (CI 95%) |  | 0.92 (0.62 - 1.00) |  |  |  |  |  |  |  |  |  |  |  |  |  |
|  | F1-score |  | 0.485 |  |  |  |  |  |  |  |  |  |  |  |  |  |
| rTransparent | Sensibility (CI 95%) |  | 0.88 (0.68 - 0.97) | 0.82 (0.65 - 0.93) | 0.11 (0 - 0.48) |  | 0.67 (0.22 - 0.96) |  |  |  |  |  |  | 1.00 (0.90 - 1.00) |  | 1.00 (0.90 - 1.00) |
|  | Specificity (CI 95%) |  | 0.92 (0.62 - 1.00) | 0.67 (0.09 - 0.99) | 1 (0.87 - 1) |  | 1.00 (0.88 - 1.00) |  |  |  |  |  |  | incalculable |  | incalculable |
|  | F1-score |  | 0.913 | 0.885 | 0.20 |  | 0.800 |  |  |  |  |  |  | 1.000 |  | 1.000 |
| Unpaywall | Sensibility (CI 95%) |  |  |  |  | 1 (0.90-1) |  |  |  |  |  |  |  |  |  |  |
|  | Specificity (CI 95%) |  |  |  |  | incalculable |  |  |  |  |  |  |  |  |  |  |
|  | F1-score |  |  |  |  | 1.00 |  |  |  |  |  |  |  |  |  |  |

### ***Sensitivity analysis of the performance of automated tools by journal: The BMJ***

| Tool | Metric | RCT | Registration | Intention to share data | Open data sharing | Open access publishing | Code sharing | Publication 1year (for RCT) | Protocol sharing | Statistical analysis plan sharing | Utilization of reporting guideline | Preprint | Author contribution | COI statement | Reporting ORCID identifiers | Funding statement |
| --- | --- | --- | --- | --- | --- | --- | --- | --- | --- | --- | --- | --- | --- | --- | --- | --- |
| CTregistries | Sensibility (CI 95%) |  | 0.64 (0.41 - 0.83) |  |  |  |  |  |  |  |  |  |  |  |  |  |
|  | Specificity (CI 95%) |  | 0.56 (0.21 - 0.86) |  |  |  |  |  |  |  |  |  |  |  |  |  |
|  | F1-score |  | 0.700 |  |  |  |  |  |  |  |  |  |  |  |  |  |
| ContriBOT | Sensibility (CI 95%) |  |  |  |  |  |  |  |  |  |  |  | 1.00 (0.89 - 1.00) |  | 0.97 (0.83 - 1.00) |  |
|  | Specificity (CI 95%) |  |  |  |  |  |  |  |  |  |  |  | incalculable |  | incalculable |  |
|  | F1-score |  |  |  |  |  |  |  |  |  |  |  | 1.000 |  | 0.984 |  |
| Data Seer | Sensibility (CI 95%) |  | 0.86 (0.65 - 0.97) | 0.07 (0.01 - 0.23) |  |  | 0.22 (0.03 - 0.60) |  |  |  |  | incalculable |  |  |  |  |
|  | Specificity (CI 95%) |  | 1.00 (0.66 - 1.00) | 1.00 (0.16 - 1.00) |  |  | 0.95 (0.77 - 1.00) |  |  |  |  | 1.00 (0.89 - 1.00) |  |  |  |  |
|  | F1-score |  | 0.927 | 0.129 |  |  | 0.333 |  |  |  |  | incalculable |  |  |  |  |
| Llama 3.70B | Sensibility (CI 95%) | 1.00 (0.69 - 1.00) | 0.77 (0.55 - 0.92) | 0.83 (0.64 - 0.94) | 0.56 (0.21 - 0.86) | 0.42 (0.25 - 0.61) | 1.00 (0.66 - 1.00) | 0.00 (0.00 - 0.60) | 0.77 (0.55 - 0.92) | 1.00 (0.29 - 1.00) | 0.92 (0.64 - 1.00) | incalculable | 0.94 (0.79 - 0.99) | 0.65 (0.45 - 0.81) | 0.13 (0.04 - 0.30) | 0.90 (0.74 - 0.98) |
|  | Specificity (CI 95%) | 0.95 (0.76 - 1.00) | 0.78 (0.40 - 0.97) | 0.50 (0.01 - 0.99) | 0.91 (0.71 - 0.99) | incalculable | 0.86 (0.65 - 0.97) | 1.00 (0.54 - 1.00) | 0.67 (0.30 - 0.93) | 0.36 (0.19 - 0.56) | 0.78 (0.52 - 0.94) | 0.97 (0.83 - 1.00) | incalculable | incalculable | incalculable | incalculable |
|  | F1-score | 0.952 | 0.829 | 0.889 | 0.63 | 0.591 | 0.857 | incalculable | 0.810 | 0.250 | 0.828 | incalculable | 0.967 | 0.784 | 0.229 | 0.949 |
| OddPub | Sensibility (CI 95%) |  |  | 0.93 (0.77 - 0.99) | 0 (0 - 0.34) |  | 0.44 (0.14 - 0.79) |  |  |  |  |  |  |  |  |  |
|  | Specificity (CI 95%) |  |  | 0.00 (0.00 - 0.84) | 1 (0.85 - 1) |  | 1.00 (0.85 - 1.00) |  |  |  |  |  |  |  |  |  |
|  | F1-score |  |  | 0.931 | incalculable |  | 0.615 |  |  |  |  |  |  |  |  |  |
| SciScore | Sensibility (CI 95%) |  | 0.09 (0.01 - 0.29) | 0.32 (0.16 - 0.52) |  |  | 0.11 (0.00 - 0.48) |  |  |  |  |  |  |  |  |  |
|  | Specificity (CI 95%) |  | 1.00 (0.63 - 1.00) | 1.00 (0.16 - 1.00) |  |  | 1.00 (0.84 - 1.00) |  |  |  |  |  |  |  |  |  |
|  | F1-score |  | 0.167 | 0.486 |  |  | 0.200 |  |  |  |  |  |  |  |  |  |
| TNRscreener | Sensibility (CI 95%) |  | 0.64 (0.41 - 0.83) |  |  |  |  |  |  |  |  |  |  |  |  |  |
|  | Specificity (CI 95%) |  | 0.56 (0.21 - 0.86) |  |  |  |  |  |  |  |  |  |  |  |  |  |
|  | F1-score |  | 0.700 |  |  |  |  |  |  |  |  |  |  |  |  |  |
| rtransparent | Sensibility (CI 95%) |  | 1.00 (0.85 - 1.00) | 0.97 (0.82 - 1.00) | 0 (0 - 0.34) |  | 0.22 (0.03 - 0.60) |  |  |  |  |  |  | 1.00 (0.89 - 1.00) |  | 1.00 (0.89 - 1.00) |
|  | Specificity (CI 95%) |  | 0.78 (0.40 - 0.97) | 0.00 (0.00 - 0.84) | 1 (0.85 - 1) |  | 1.00 (0.85 - 1.00) |  |  |  |  |  |  | incalculable |  | incalculable |
|  | F1-score |  | 0.957 | 0.949 | incalculable |  | 0.364 |  |  |  |  |  |  | 1.000 |  | 1.000 |
| Unpaywall | Sensibility (CI 95%) |  |  |  |  | 1 (0.89-1) |  |  |  |  |  |  |  |  |  |  |
|  | Specificity (CI 95%) |  |  |  |  | incalculable |  |  |  |  |  |  |  |  |  |  |
|  | F1-score |  |  |  |  | 1.00 |  |  |  |  |  |  |  |  |  |  |

### ***Sensitivity analysis of the performance of automated tools by journal: CMAJ***

| Tool | Metric | RCT | Registration | Intention to share data | Open data sharing | Open access publishing | Code sharing | Publication 1year (for RCT) | Protocol sharing | Statistical analysis plan sharing | Utilization of reporting guideline | Preprint | Author contribution | COI statement | Reporting ORCID identifiers | Funding statement |
| --- | --- | --- | --- | --- | --- | --- | --- | --- | --- | --- | --- | --- | --- | --- | --- | --- |
| CTregistries | Sensibility (CI 95%) |  | 0.50 (0.12 - 0.88) |  |  |  |  |  |  |  |  |  |  |  |  |  |
|  | Specificity (CI 95%) |  | 1.00 (0.72 - 1.00) |  |  |  |  |  |  |  |  |  |  |  |  |  |
|  | F1-score |  | 0.667 |  |  |  |  |  |  |  |  |  |  |  |  |  |
| ContriBOT | Sensibility (CI 95%) |  |  |  |  |  |  |  |  |  |  |  | 1.00 (0.79 - 1.00) |  | incalculable |  |
|  | Specificity (CI 95%) |  |  |  |  |  |  |  |  |  |  |  | 1.00 (0.03 - 1.00) |  | 1.00 (0.80 - 1.00) |  |
|  | F1-score |  |  |  |  |  |  |  |  |  |  |  | 1.000 |  | incalculable |  |
| Data Seer | Sensibility (CI 95%) |  | 1.00 (0.40 - 1.00) | 0.09 (0.00 - 0.41) |  |  | 0.00 (0.00 - 0.84) |  |  |  |  | incalculable |  |  |  |  |
|  | Specificity (CI 95%) |  | 1.00 (0.72 - 1.00) | 1.00 (0.40 - 1.00) |  |  | 1.00 (0.75 - 1.00) |  |  |  |  | 1.00 (0.78 - 1.00) |  |  |  |  |
|  | F1-score |  | 1.000 | 0.167 |  |  | incalculable |  |  |  |  | incalculable |  |  |  |  |
| Llama 3.3 70B | Sensibility (CI 95%) | 1.00 (0.29 - 1.00) | 0.67 (0.22 - 0.96) | 0.85 (0.55 - 0.98) | 1 (0.03 - 1) | 0.59 (0.33 - 0.82) | 0.67 (0.09 - 0.99) | 0.00 (0.00 - 0.97) | 0.75 (0.19 - 0.99) | incalculable | 0.60 (0.15 - 0.95) | incalculable | 0.88 (0.62 - 0.98) | 0.59 (0.33 - 0.82) | incalculable | 0.82 (0.57 - 0.96) |
|  | Specificity (CI 95%) | 1.00 (0.77 - 1.00) | 1.00 (0.72 - 1.00) | 0.00 (0.00 - 0.60) | 0.81 (0.54 - 0.96) | incalculable | 0.93 (0.66 - 1.00) | 0.00 (0.00 - 0.84) | 0.15 (0.02 - 0.45) | 0.24 (0.07 - 0.50) | 1.00 (0.74 - 1.00) | 0.94 (0.71 - 1.00) | 0.00 (0.00 - 0.97) | incalculable | 1.00 (0.80 - 1.00) | incalculable |
|  | F1-score | 1.000 | 0.800 | 0.786 | 0.40 | 0.741 | 0.667 | incalculable | 0.333 | incalculable | 0.750 | incalculable | 0.903 | 0.741 | incalculable | 0.903 |
| ODDPub | Sensibility (CI 95%) |  |  | 0.31 (0.09 - 0.61) | 1 (0.03 - 1) |  | 0.00 (0.00 - 0.71) |  |  |  |  |  |  |  |  |  |
|  | Specificity (CI 95%) |  |  | 1.00 (0.40 - 1.00) | 0.94 (0.7 - 1) |  | 1.00 (0.77 - 1.00) |  |  |  |  |  |  |  |  |  |
|  | F1-score |  |  | 0.471 | 0.67 |  | incalculable |  |  |  |  |  |  |  |  |  |
| SciScore | Sensibility (CI 95%) |  | 0.17 (0.00 - 0.64) | 0.36 (0.11 - 0.69) |  |  | 0.00 (0.00 - 0.84) |  |  |  |  |  |  |  |  |  |
|  | Specificity (CI 95%) |  | 1.00 (0.63 - 1.00) | 0.33 (0.01 - 0.91) |  |  | 1.00 (0.74 - 1.00) |  |  |  |  |  |  |  |  |  |
|  | F1-score |  | 0.286 | 0.471 |  |  | incalculable |  |  |  |  |  |  |  |  |  |
| TNRscreener | Sensibility (CI 95%) |  | 0.67 (0.22 - 0.96) |  |  |  |  |  |  |  |  |  |  |  |  |  |
|  | Specificity (CI 95%) |  | 1.00 (0.72 - 1.00) |  |  |  |  |  |  |  |  |  |  |  |  |  |
|  | F1-score |  | 0.800 |  |  |  |  |  |  |  |  |  |  |  |  |  |
| rTransparent | Sensibility (CI 95%) |  | 0.83 (0.36 - 1.00) | 0.77 (0.46 - 0.95) | 0 (0 - 0.98) |  | 0.00 (0.00 - 0.71) |  |  |  |  |  |  | 1.00 (0.80 - 1.00) |  | 1.00 (0.80 - 1.00) |
|  | Specificity (CI 95%) |  | 1.00 (0.72 - 1.00) | 0.75 (0.19 - 0.99) | 0.94 (0.7 - 1) |  | 1.00 (0.77 - 1.00) |  |  |  |  |  |  | incalculable |  | incalculable |
|  | F1-score |  | 0.909 | 0.833 | incalculable |  | incalculable |  |  |  |  |  |  | 1.000 |  | 1.000 |
| Unpaywall | Sensibility (CI 95%) |  |  |  |  | 1 (0.80-1) |  |  |  |  |  |  |  |  |  |  |
|  | Specificity (CI 95%) |  |  |  |  | incalculable |  |  |  |  |  |  |  |  |  |  |
|  | F1-score |  |  |  |  | 1.00 |  |  |  |  |  |  |  |  |  |  |

### ***Sensitivity analysis of the performance of automated tools by journal: JAMA***

| Tool | Metric | RCT | Registration | Intention to share data | Open data sharing | Open access publishing | Code sharing | Publication 1year (for RCT) | Protocol sharing | Statistical analysis plan sharing | Utilization of reporting guideline | Preprint | Author contribution | COI statement | Reporting ORCID identifiers | Funding statement |
| --- | --- | --- | --- | --- | --- | --- | --- | --- | --- | --- | --- | --- | --- | --- | --- | --- |
| CTregistries | Sensibility (CI 95%) |  | 0.38 (0.15 - 0.65) |  |  |  |  |  |  |  |  |  |  |  |  |  |
|  | Specificity (CI 95%) |  | 0.88 (0.62 - 0.98) |  |  |  |  |  |  |  |  |  |  |  |  |  |
|  | F1-score |  | 0.500 |  |  |  |  |  |  |  |  |  |  |  |  |  |
| ContriBOT | Sensibility (CI 95%) |  |  |  |  |  |  |  |  |  |  |  | 0.91 (0.75 - 0.98) |  | incalculable |  |
|  | Specificity (CI 95%) |  |  |  |  |  |  |  |  |  |  |  | incalculable |  | 1.00 (0.89 - 1.00) |  |
|  | F1-score |  |  |  |  |  |  |  |  |  |  |  | 0.951 |  | incalculable |  |
| Data Seer | Sensibility (CI 95%) |  | 0.44 (0.20 - 0.70) | 0.00 (0.00 - 0.22) |  |  | 0.00 (0.00 - 0.60) |  |  |  |  | incalculable |  |  |  |  |
|  | Specificity (CI 95%) |  | 1.00 (0.75 - 1.00) | 1.00 (0.77 - 1.00) |  |  | 1.00 (0.86 - 1.00) |  |  |  |  | 1.00 (0.88 - 1.00) |  |  |  |  |
|  | F1-score |  | 0.609 | incalculable |  |  | incalculable |  |  |  |  | incalculable |  |  |  |  |
| Llama 3.3 70B | Sensibility (CI 95%) | 1.00 (0.72 - 1.00) | 0.69 (0.41 - 0.89) | 0.62 (0.35 - 0.85) | 0.33 (0.01 - 0.91) | 0.09 (0.02 - 0.25) | 0.50 (0.07 - 0.93) | 0.60 (0.15 - 0.95) | 0.62 (0.35 - 0.85) | 0.89 (0.52 - 1.00) | 0.56 (0.21 - 0.86) | incalculable | 0.78 (0.60 - 0.91) | 0.78 (0.60 - 0.91) | incalculable | 0.70 (0.51 - 0.85) |
|  | Specificity (CI 95%) | 0.90 (0.70 - 0.99) | 1.00 (0.79 - 1.00) | 0.75 (0.48 - 0.93) | 1 (0.88 - 1) | incalculable | 0.75 (0.55 - 0.89) | 0.00 (0.00 - 0.46) | 0.69 (0.41 - 0.89) | 0.65 (0.43 - 0.84) | 0.87 (0.66 - 0.97) | 1.00 (0.89 - 1.00) | incalculable | incalculable | 1.00 (0.89 - 1.00) | 1.00 (0.16 - 1.00) |
|  | F1-score | 0.917 | 0.815 | 0.667 | 0.50 | 0.171 | 0.308 | 0.429 | 0.645 | 0.640 | 0.588 | incalculable | 0.877 | 0.877 | incalculable | 0.824 |
| ODDPub | Sensibility (CI 95%) |  |  | 0.69 (0.41 - 0.89) | 0.33 (0.01 - 0.91) |  | 0.00 (0.00 - 0.60) |  |  |  |  |  |  |  |  |  |
|  | Specificity (CI 95%) |  |  | 0.88 (0.62 - 0.98) | 1 (0.88 - 1) |  | 0.93 (0.76 - 0.99) |  |  |  |  |  |  |  |  |  |
|  | F1-score |  |  | 0.759 | 0.5 |  | incalculable |  |  |  |  |  |  |  |  |  |
| SciScore | Sensibility (CI 95%) |  | 0.00 (0.00 - 0.25) | 0.00 (0.00 - 0.28) |  |  | 0.00 (0.00 - 0.71) |  |  |  |  |  |  |  |  |  |
|  | Specificity (CI 95%) |  | 1.00 (0.66 - 1.00) | 0.73 (0.39 - 0.94) |  |  | 1.00 (0.82 - 1.00) |  |  |  |  |  |  |  |  |  |
|  | F1-score |  | incalculable |  |  |  | incalculable |  |  |  |  |  |  |  |  |  |
| TNRscreener | Sensibility (CI 95%) |  | 0.38 (0.15 - 0.65) |  |  |  |  |  |  |  |  |  |  |  |  |  |
|  | Specificity (CI 95%) |  | 0.88 (0.62 - 0.98) |  |  |  |  |  |  |  |  |  |  |  |  |  |
|  | F1-score |  | 0.500 |  |  |  |  |  |  |  |  |  |  |  |  |  |
| rTransparent | Sensibility (CI 95%) |  | 0.88 (0.62 - 0.98) | 0.69 (0.41 - 0.89) | 0.33 (0.01 - 0.91) |  | 0.00 (0.00 - 0.60) |  |  |  |  |  |  | 1.00 (0.89 - 1.00) |  | 1.00 (0.88 - 1.00) |
|  | Specificity (CI 95%) |  | 0.94 (0.70 - 1.00) | 0.81 (0.54 - 0.96) | 0.97 (0.82 - 1) |  | 0.93 (0.76 - 0.99) |  |  |  |  |  |  | incalculable |  | 1.00 (0.16 - 1.00) |
|  | F1-score |  | 0.903 | 0.733 | 0.40 |  | incalculable |  |  |  |  |  |  | 1.000 |  | 1.000 |
| Unpaywall | Sensibility (CI 95%) |  |  |  |  | 0.90 (0.75-0.98) |  |  |  |  |  |  |  |  |  |  |
|  | Specificity (CI 95%) |  |  |  |  | incalculable |  |  |  |  |  |  |  |  |  |  |
|  | F1-score |  |  |  |  | 0.95 |  |  |  |  |  |  |  |  |  |  |

### ***Sensitivity analysis of the performance of automated tools by journal: JAMA Network Open***

| Tool | Metric | RCT | Registration | Intention to share data | Open data sharing | Open access publishing | Code sharing | Publication 1year (for RCT) | Protocol sharing | Statistical analysis plan sharing | Utilization of reporting guideline | Preprint | Author contribution | COI statement | Reporting ORCID identifiers | Funding statement |
| --- | --- | --- | --- | --- | --- | --- | --- | --- | --- | --- | --- | --- | --- | --- | --- | --- |
| CTregistries | Sensibility (CI 95%) |  | 0.35 (0.16 - 0.57) |  |  |  |  |  |  |  |  |  |  |  |  |  |
|  | Specificity (CI 95%) |  | 0.90 (0.68 - 0.99) |  |  |  |  |  |  |  |  |  |  |  |  |  |
|  | F1-score |  | 0.485 |  |  |  |  |  |  |  |  |  |  |  |  |  |
| ContriBOT | Sensibility (CI 95%) |  |  |  |  |  |  |  |  |  |  |  | 1.00 (0.92 - 1.00) |  | incalculable |  |
|  | Specificity (CI 95%) |  |  |  |  |  |  |  |  |  |  |  | incalculable |  | 1.00 (0.92 - 1.00) |  |
|  | F1-score |  |  |  |  |  |  |  |  |  |  |  | 1.000 |  | incalculable |  |
| Data Seer | Sensibility (CI 95%) |  | 0.87 (0.66 - 0.97) | 0.88 (0.68 - 0.97) |  |  | 0.00 (0.00 - 0.71) |  |  |  |  | incalculable |  |  |  |  |
|  | Specificity (CI 95%) |  | 0.80 (0.56 - 0.94) | 0.63 (0.38 - 0.84) |  |  | 1.00 (0.91 - 1.00) |  |  |  |  | 1.00 (0.92 - 1.00) |  |  |  |  |
|  | F1-score |  | 0.851 | 0.808 |  |  | incalculable |  |  |  |  | incalculable |  |  |  |  |
| Llama 3.3 70B | Sensibility (CI 95%) | 1.00 (0.75 - 1.00) | 0.70 (0.47 - 0.87) | 0.92 (0.73 - 0.99) | 0 (0 - 0.98) | 0.51 (0.35 - 0.67) | 0.67 (0.09 - 0.99) | 1.00 (0.40 - 1.00) | 0.76 (0.55 - 0.91) | 0.80 (0.28 - 0.99) | 0.74 (0.58 - 0.86) | incalculable | 0.74 (0.59 - 0.86) | 0.77 (0.61 - 0.88) | incalculable | 0.87 (0.73 - 0.96) |
|  | Specificity (CI 95%) | 0.77 (0.58 - 0.90) | 0.80 (0.56 - 0.94) | 0.47 (0.24 - 0.71) | 0.9 (0.77 - 0.97) | incalculable | 0.47 (0.32 - 0.64) | 0.21 (0.05 - 0.51) | 0.44 (0.22 - 0.69) | 0.34 (0.20 - 0.51) | 0.00 (0.00 - 0.97) | 1.00 (0.92 - 1.00) | incalculable | incalculable | 1.00 (0.92 - 1.00) | 0.50 (0.07 - 0.93) |
|  | F1-score | 0.788 | 0.744 | 0.786 | incalculable | 0.677 | 0.154 | 0.421 | 0.704 | 0.235 | 0.838 | incalculable | 0.853 | 0.868 | incalculable | 0.907 |
| ODDPub | Sensibility (CI 95%) |  |  | 0.71 (0.49 - 0.87) | 0 (0 - 0.98) |  | 0.00 (0.00 - 0.71) |  |  |  |  |  |  |  |  |  |
|  | Specificity (CI 95%) |  |  | 0.79 (0.54 - 0.94) | 1 (0.92 - 1) |  | 0.93 (0.80 - 0.98) |  |  |  |  |  |  |  |  |  |
|  | F1-score |  |  | 0.756 | incalculable |  | incalculable |  |  |  |  |  |  |  |  |  |
| SciScore | Sensibility (CI 95%) |  | 0.00 (0.00 - 0.18) | 0.19 (0.05 - 0.42) |  |  | 0.00 (0.00 - 0.71) |  |  |  |  |  |  |  |  |  |
|  | Specificity (CI 95%) |  | 1.00 (0.79 - 1.00) | 0.79 (0.49 - 0.95) |  |  | 1.00 (0.89 - 1.00) |  |  |  |  |  |  |  |  |  |
|  | F1-score |  | incalculable | 0.286 |  |  | incalculable |  |  |  |  |  |  |  |  |  |
| TNRscreener | Sensibility (CI 95%) |  | 0.35 (0.16 - 0.57) |  |  |  |  |  |  |  |  |  |  |  |  |  |
|  | Specificity (CI 95%) |  | 0.90 (0.68 - 0.99) |  |  |  |  |  |  |  |  |  |  |  |  |  |
|  | F1-score |  | 0.485 |  |  |  |  |  |  |  |  |  |  |  |  |  |
| rTransparent | Sensibility (CI 95%) |  | 0.96 (0.78 - 1.00) | 0.71 (0.49 - 0.87) | 0 (0 - 0.98) |  | 0.00 (0.00 - 0.71) |  |  |  |  |  |  | 1.00 (0.92 - 1.00) |  | 0.92 (0.79 - 0.98) |
|  | Specificity (CI 95%) |  | 0.80 (0.56 - 0.94) | 0.79 (0.54 - 0.94) | 1 (0.92 - 1) |  | 0.95 (0.83 - 0.99) |  |  |  |  |  |  | incalculable |  | 0.50 (0.07 - 0.93) |
|  | F1-score |  | 0.898 | 0.756 | incalculable |  | incalculable |  |  |  |  |  |  | 1.000 |  | 0.935 |
| Unpaywall | Sensibility (CI 95%) |  |  |  |  | 1 (0.91-1) |  |  |  |  |  |  |  |  |  |  |
|  | Specificity (CI 95%) |  |  |  |  | incalculable |  |  |  |  |  |  |  |  |  |  |
|  | F1-score |  |  |  |  | 1 |  |  |  |  |  |  |  |  |  |  |

### ***Sensitivity analysis of the performance of automated tools by journal: Lancet***

| Tool | Metric | RCT | Registration | Intention to share data | Open data sharing | Open access publishing | Code sharing | Publication 1year (for RCT) | Protocol sharing | Statistical analysis plan sharing | Utilization of reporting guideline | Preprint | Author contribution | COI statement | Reporting ORCID identifiers | Funding statement |
| --- | --- | --- | --- | --- | --- | --- | --- | --- | --- | --- | --- | --- | --- | --- | --- | --- |
| CTregistries | Sensibility (CI 95%) |  | 0.73 (0.50 - 0.89) |  |  |  |  |  |  |  |  |  |  |  |  |  |
|  | Specificity (CI 95%) |  | 0.83 (0.52 - 0.98) |  |  |  |  |  |  |  |  |  |  |  |  |  |
|  | F1-score |  | 0.800 |  |  |  |  |  |  |  |  |  |  |  |  |  |
| ContriBOT | Sensibility (CI 95%) |  |  |  |  |  |  |  |  |  |  |  | 0.00 (0.00 - 0.10) |  | incalculable |  |
|  | Specificity (CI 95%) |  |  |  |  |  |  |  |  |  |  |  | incalculable |  | 1.00 (0.90 - 1.00) |  |
|  | F1-score |  |  |  |  |  |  |  |  |  |  |  | incalculable |  | incalculable |  |
| Llama 3.3 70B | Sensibility (CI 95%) | 0.42 (0.15 - 0.72) | 1.00 (0.85 - 1.00) | 1.00 (0.89 - 1.00) | 0.86 (0.42 - 1) | 0.94 (0.71 - 1.00) | 0.40 (0.05 - 0.85) | 0.00 (0.00 - 0.41) | 0.92 (0.73 - 0.99) | 1.00 (0.75 - 1.00) | 0.93 (0.66 - 1.00) | incalculable | 0.91 (0.76 - 0.98) | 1.00 (0.90 - 1.00) | incalculable | 0.91 (0.76 - 0.98) |
|  | Specificity (CI 95%) | 1.00 (0.85 - 1.00) | 0.92 (0.62 - 1.00) | 0.00 (0.00 - 0.71) | 1 (0.87 - 1) | 0.76 (0.50 - 0.93) | 0.69 (0.49 - 0.85) | 1.00 (0.48 - 1.00) | 0.30 (0.07 - 0.65) | 0.10 (0.01 - 0.30) | 0.90 (0.68 - 0.99) | 0.85 (0.69 - 0.95) | incalculable | incalculable | 1.00 (0.90 - 1.00) | incalculable |
|  | F1-score | 0.588 | 0.978 | 0.954 | 0.92 | 0.865 | 0.250 | incalculable | 0.830 | 0.578 | 0.897 | incalculable | 0.954 | 1.000 | incalculable | 0.954 |
| ODDPub | Sensibility (CI 95%) |  |  | 0.81 (0.63 - 0.93) | 0.43 (0.1 - 0.82) |  | 0.80 (0.28 - 0.99) |  |  |  |  |  |  |  |  |  |
|  | Specificity (CI 95%) |  |  | 0.33 (0.01 - 0.91) | 0.96 (0.81 - 1) |  | 0.76 (0.56 - 0.90) |  |  |  |  |  |  |  |  |  |
|  | F1-score |  |  | 0.862 | 0.55 |  | 0.500 |  |  |  |  |  |  |  |  |  |
| TNRscreener | Sensibility (CI 95%) |  | 0.73 (0.50 - 0.89) |  |  |  |  |  |  |  |  |  |  |  |  |  |
|  | Specificity (CI 95%) |  | 0.83 (0.52 - 0.98) |  |  |  |  |  |  |  |  |  |  |  |  |  |
|  | F1-score |  | 0.800 |  |  |  |  |  |  |  |  |  |  |  |  |  |
| rTransparent | Sensibility (CI 95%) |  | 0.00 (0.00 - 0.15) | 0.81 (0.63 - 0.93) | 0.43 (0.1 - 0.82) |  | 0.60 (0.15 - 0.95) |  |  |  |  |  |  | 1.00 (0.90 - 1.00) |  | 1.00 (0.90 - 1.00) |
|  | Specificity (CI 95%) |  | 1.00 (0.74 - 1.00) | 0.33 (0.01 - 0.91) | 0.96 (0.81 - 1) |  | 0.86 (0.68 - 0.96) |  |  |  |  |  |  | incalculable |  | incalculable |
|  | F1-score |  | incalculable | 0.862 | 0.55 |  | 0.500 |  |  |  |  |  |  | 1.000 |  | 1.000 |
| Unpaywall | Sensibility (CI 95%) |  |  |  |  | incalculable |  |  |  |  |  |  |  |  |  |  |
|  | Specificity (CI 95%) |  |  |  |  | incalculable |  |  |  |  |  |  |  |  |  |  |
|  | F1-score |  |  |  |  | incalculable |  |  |  |  |  |  |  |  |  |  |

### ***Sensitivity analysis of the performance of automated tools by journal: NEJM***

| Tool | Metric | RCT | Registration | Intention to share data | Open data sharing | Open access publishing | Code sharing | Publication 1year (for RCT) | Protocol sharing | Statistical analysis plan sharing | Utilization of reporting guideline | Preprint | Author contribution | COI statement | Reporting ORCID identifiers | Funding statement |
| --- | --- | --- | --- | --- | --- | --- | --- | --- | --- | --- | --- | --- | --- | --- | --- | --- |
| CTregistries | Sensibility (CI 95%) |  | 0.83 (0.59 - 0.96) |  |  |  |  |  |  |  |  |  |  |  |  |  |
|  | Specificity (CI 95%) |  | 0.80 (0.28 - 0.99) |  |  |  |  |  |  |  |  |  |  |  |  |  |
|  | F1-score |  | 0.882 |  |  |  |  |  |  |  |  |  |  |  |  |  |
| ContriBOT | Sensibility (CI 95%) |  |  |  |  |  |  |  |  |  |  |  | 0.67 (0.09 - 0.99) |  | 0.00 (0.00 - 0.20) |  |
|  | Specificity (CI 95%) |  |  |  |  |  |  |  |  |  |  |  | 0.85 (0.62 - 0.97) |  | 1.00 (0.54 - 1.00) |  |
|  | F1-score |  |  |  |  |  |  |  |  |  |  |  | 0.500 |  | incalculable |  |
| Data Seer | Sensibility (CI 95%) |  | 0.61 (0.36 - 0.83) | 0.00 (0.00 - 0.28) |  |  | incalculable |  |  |  |  | incalculable |  |  |  |  |
|  | Specificity (CI 95%) |  | 0.80 (0.28 - 0.99) | 1.00 (0.74 - 1.00) |  |  | 1.00 (0.85 - 1.00) |  |  |  |  | 1.00 (0.85 - 1.00) |  |  |  |  |
|  | F1-score |  | 0.733 | incalculable |  |  | incalculable |  |  |  |  | incalculable |  |  |  |  |
| Llama 3.3 70B | Sensibility (CI 95%) | 1.00 (0.77 - 1.00) | 0.67 (0.41 - 0.87) | 0.91 (0.59 - 1.00) | incalculable | 0.00 (0.00 - 0.15) | incalculable | 0.89 (0.52 - 1.00) | 1.00 (0.81 - 1.00) | 1.00 (0.81 - 1.00) | 0.00 (0.00 - 0.97) | incalculable | 1.00 (0.29 - 1.00) | 0.87 (0.66 - 0.97) | 0.00 (0.00 - 0.20) | 1.00 (0.85 - 1.00) |
|  | Specificity (CI 95%) | 0.67 (0.30 - 0.93) | 1.00 (0.48 - 1.00) | 0.50 (0.21 - 0.79) | incalculable | incalculable | 0.39 (0.20 - 0.61) | 0.00 (0.00 - 0.52) | 1.00 (0.48 - 1.00) | 0.60 (0.15 - 0.95) | 0.82 (0.60 - 0.95) | 1.00 (0.85 - 1.00) | 0.05 (0.00 - 0.25) | incalculable | 1.00 (0.54 - 1.00) | incalculable |
|  | F1-score | 0.903 | 0.800 | 0.741 | incalculable | incalculable | incalculable | 0.727 | 1.000 | 0.947 | incalculable | incalculable | 0.240 | 0.930 | incalculable | 1.000 |
| ODDPub | Sensibility (CI 95%) |  |  | 1.00 (0.72 - 1.00) | incalculable |  | incalculable |  |  |  |  |  |  |  |  |  |
|  | Specificity (CI 95%) |  |  | 0.00 (0.00 - 0.26) | incalculable |  | 0.65 (0.43 - 0.84) |  |  |  |  |  |  |  |  |  |
|  | F1-score |  |  | 0.647 | incalculable |  | incalculable |  |  |  |  |  |  |  |  |  |
| SciScore | Sensibility (CI 95%) |  | 0.08 (0.00 - 0.36) | 0.11 (0.00 - 0.48) |  |  | incalculable |  |  |  |  |  |  |  |  |  |
|  | Specificity (CI 95%) |  | 1.00 (0.29 - 1.00) | 1.00 (0.59 - 1.00) |  |  | 1.00 (0.79 - 1.00) |  |  |  |  |  |  |  |  |  |
|  | F1-score |  | 0.143 | 0.200 |  |  | incalculable |  |  |  |  |  |  |  |  |  |
| TNRscreener | Sensibility (CI 95%) |  | 0.83 (0.59 - 0.96) |  |  |  |  |  |  |  |  |  |  |  |  |  |
|  | Specificity (CI 95%) |  | 0.80 (0.28 - 0.99) |  |  |  |  |  |  |  |  |  |  |  |  |  |
|  | F1-score |  | 0.882 |  |  |  |  |  |  |  |  |  |  |  |  |  |
| rTransparent | Sensibility (CI 95%) |  | 0.89 (0.65 - 0.99) | 1.00 (0.72 - 1.00) | incalculable |  | incalculable |  |  |  |  |  |  | 1.00 (0.85 - 1.00) |  | 1.00 (0.85 - 1.00) |
|  | Specificity (CI 95%) |  | 1.00 (0.48 - 1.00) | 0.00 (0.00 - 0.26) | incalculable |  | 0.96 (0.78 - 1.00) |  |  |  |  |  |  | incalculable |  | incalculable |
|  | F1-score |  | 0.941 | 0.647 | incalculable |  | incalculable |  |  |  |  |  |  | 1.000 |  | 1.000 |
| Unpaywall | Sensibility (CI 95%) |  |  |  |  | incalculable |  |  |  |  |  |  |  |  |  |  |
|  | Specificity (CI 95%) |  |  |  |  | incalculable |  |  |  |  |  |  |  |  |  |  |
|  | F1-score |  |  |  |  | incalculable |  |  |  |  |  |  |  |  |  |  |

### ***Sensitivity analysis of the performance of automated tools by journal: Nature Medicine***

| Tool | Metric | RCT | Registration | Intention to share data | Open data sharing | Open access publishing | Code sharing | Publication 1year (for RCT) | Protocol sharing | Statistical analysis plan sharing | Utilization of reporting guideline | Preprint | Author contribution | COI statement | Reporting ORCID identifiers | Funding statement |
| --- | --- | --- | --- | --- | --- | --- | --- | --- | --- | --- | --- | --- | --- | --- | --- | --- |
| CTregistries | Sensibility (CI 95%) |  | 0.89 (0.67 - 0.99) |  |  |  |  |  |  |  |  |  |  |  |  |  |
|  | Specificity (CI 95%) |  | 0.77 (0.46 - 0.95) |  |  |  |  |  |  |  |  |  |  |  |  |  |
|  | F1-score |  | 0.872 |  |  |  |  |  |  |  |  |  |  |  |  |  |
| ContriBOT | Sensibility (CI 95%) |  |  |  |  |  |  |  |  |  |  |  | 0.97 (0.84 - 1.00) |  | 1.00 (0.88 - 1.00) |  |
|  | Specificity (CI 95%) |  |  |  |  |  |  |  |  |  |  |  | incalculable |  | 0.00 (0.00 - 0.84) |  |
|  | F1-score |  |  |  |  |  |  |  |  |  |  |  | 0.984 |  | 0.968 |  |
| Data Seer | Sensibility (CI 95%) |  | 0.88 (0.62 - 0.98) | 0.88 (0.69 - 0.97) |  |  | 0.43 (0.18 - 0.71) |  |  |  |  | incalculable |  |  |  |  |
|  | Specificity (CI 95%) |  | 0.90 (0.55 - 1.00) | 0.00 (0.00 - 0.97) |  |  | 0.92 (0.62 - 1.00) |  |  |  |  | 1.00 (0.87 - 1.00) |  |  |  |  |
|  | F1-score |  | 0.903 | 0.917 |  |  | 0.571 |  |  |  |  | incalculable |  |  |  |  |
| Llama 3.3 70B | Sensibility (CI 95%) | 1.00 (0.69 - 1.00) | 0.63 (0.38 - 0.84) | 0.93 (0.78 - 0.99) | 0.33 (0.04 - 0.78) | 0.24 (0.08 - 0.47) | 0.95 (0.74 - 1.00) | 0.50 (0.01 - 0.99) | 0.93 (0.66 - 1.00) | 1.00 (0.48 - 1.00) | 0.80 (0.28 - 0.99) | incalculable | 0.94 (0.79 - 0.99) | 0.94 (0.79 - 0.99) | 0.00 (0.00 - 0.12) | 0.81 (0.64 - 0.93) |
|  | Specificity (CI 95%) | 0.82 (0.60 - 0.95) | 0.92 (0.64 - 1.00) | 0.00 (0.00 - 0.84) | 0.85 (0.65 - 0.96) | 0.91 (0.59 - 1.00) | 0.69 (0.39 - 0.91) | 1.00 (0.66 - 1.00) | 0.11 (0.01 - 0.35) | 0.19 (0.06 - 0.38) | 0.59 (0.39 - 0.78) | 1.00 (0.89 - 1.00) | incalculable | incalculable | 1.00 (0.16 - 1.00) | incalculable |
|  | F1-score | 0.833 | 0.750 | 0.933 | 0.33 | 0.370 | 0.878 | 0.667 | 0.605 | 0.312 | 0.400 | incalculable | 0.968 | 0.968 | incalculable | 0.897 |
| ODDPub | Sensibility (CI 95%) |  |  | 0.93 (0.78 - 0.99) | 0.67 (0.22 - 0.96) |  | 0.68 (0.43 - 0.87) |  |  |  |  |  |  |  |  |  |
|  | Specificity (CI 95%) |  |  | 0.50 (0.01 - 0.99) | 0.81 (0.61 - 0.93) |  | 0.77 (0.46 - 0.95) |  |  |  |  |  |  |  |  |  |
|  | F1-score |  |  | 0.949 | 0.53 |  | 0.743 |  |  |  |  |  |  |  |  |  |
| SciScore | Sensibility (CI 95%) |  | 0.33 (0.13 - 0.59) | 0.48 (0.29 - 0.67) |  |  | 0.22 (0.06 - 0.48) |  |  |  |  |  |  |  |  |  |
|  | Specificity (CI 95%) |  | 0.92 (0.64 - 1.00) | 0.50 (0.01 - 0.99) |  |  | 1.00 (0.75 - 1.00) |  |  |  |  |  |  |  |  |  |
|  | F1-score |  | 0.480 | 0.636 |  |  | 0.364 |  |  |  |  |  |  |  |  |  |
| TNRscreener | Sensibility (CI 95%) |  | 0.89 (0.67 - 0.99) |  |  |  |  |  |  |  |  |  |  |  |  |  |
|  | Specificity (CI 95%) |  | 0.77 (0.46 - 0.95) |  |  |  |  |  |  |  |  |  |  |  |  |  |
|  | F1-score |  | 0.872 |  |  |  |  |  |  |  |  |  |  |  |  |  |
| rTransparent | Sensibility (CI 95%) |  | 0.79 (0.54 - 0.94) | 0.93 (0.78 - 0.99) | 0.67 (0.22 - 0.96) |  | 0.63 (0.38 - 0.84) |  |  |  |  |  |  | 1.00 (0.89 - 1.00) |  | 0.97 (0.84 - 1.00) |
|  | Specificity (CI 95%) |  | 1.00 (0.75 - 1.00) | 0.50 (0.01 - 0.99) | 0.81 (0.61 - 0.93) |  | 1.00 (0.75 - 1.00) |  |  |  |  |  |  | incalculable |  | incalculable |
|  | F1-score |  | 0.882 | 0.949 | 0.53 |  | 0.774 |  |  |  |  |  |  | 1.000 |  | 0.984 |
| Unpaywall | Sensibility (CI 95%) |  |  |  |  | 1 (0.83-1) |  |  |  |  |  |  |  |  |  |  |
|  | Specificity (CI 95%) |  |  |  |  | 0.36 (0.11-069) |  |  |  |  |  |  |  |  |  |  |
|  | F1-score |  |  |  |  | 0.86 |  |  |  |  |  |  |  |  |  |  |

### ***Sensitivity analysis of the performance of automated tools by journal: PLoS Medicine***

| Tool | Metric | RCT | Registration | Intention to share data | Open data sharing | Open access publishing | Code sharing | Publication 1year (for RCT) | Protocol sharing | Statistical analysis plan sharing | Utilization of reporting guideline | Preprint | Author contribution | COI statement | Reporting ORCID identifiers | Funding statement |
| --- | --- | --- | --- | --- | --- | --- | --- | --- | --- | --- | --- | --- | --- | --- | --- | --- |
| CTregistries | Sensibility (CI 95%) |  | 0.43 (0.22 - 0.66) |  |  |  |  |  |  |  |  |  |  |  |  |  |
|  | Specificity (CI 95%) |  | 0.77 (0.46 - 0.95) |  |  |  |  |  |  |  |  |  |  |  |  |  |
|  | F1-score |  | 0.545 |  |  |  |  |  |  |  |  |  |  |  |  |  |
| ContriBOT | Sensibility (CI 95%) |  |  |  |  |  |  |  |  |  |  |  | 1.00 (0.90 - 1.00) |  | 1.00 (0.90 - 1.00) |  |
|  | Specificity (CI 95%) |  |  |  |  |  |  |  |  |  |  |  | incalculable |  | incalculable |  |
|  | F1-score |  |  |  |  |  |  |  |  |  |  |  | 1.000 |  | 1.000 |  |
| Data Seer | Sensibility (CI 95%) |  | 0.90 (0.68 - 0.99) | 0.55 (0.36 - 0.72) |  |  | 0.25 (0.03 - 0.65) |  |  |  |  | incalculable |  |  |  |  |
|  | Specificity (CI 95%) |  | 0.92 (0.64 - 1.00) | incalculable |  |  | 1.00 (0.86 - 1.00) |  |  |  |  | 1.00 (0.89 - 1.00) |  |  |  |  |
|  | F1-score |  | 0.923 | 0.706 |  |  | 0.400 |  |  |  |  | incalculable |  |  |  |  |
| Llama 3.3 70B | Sensibility (CI 95%) | 1.00 (0.66 - 1.00) | 0.71 (0.48 - 0.89) | 0.68 (0.49 - 0.83) | 0.31 (0.11 - 0.59) | 0.21 (0.09 - 0.38) | 0.75 (0.35 - 0.97) | 0.00 (0.00 - 0.97) | 0.63 (0.38 - 0.84) | 1.00 (0.40 - 1.00) | 0.71 (0.53 - 0.85) | incalculable | 0.62 (0.44 - 0.78) | 0.29 (0.15 - 0.47) | 0.00 (0.00 - 0.10) | 0.85 (0.69 - 0.95) |
|  | Specificity (CI 95%) | 0.96 (0.80 - 1.00) | 0.92 (0.64 - 1.00) | incalculable | 0.83 (0.59 - 0.96) | incalculable | 0.92 (0.75 - 0.99) | 0.89 (0.52 - 1.00) | 0.40 (0.16 - 0.68) | 0.53 (0.34 - 0.72) | incalculable | 0.97 (0.85 - 1.00) | incalculable | incalculable | incalculable | incalculable |
|  | F1-score | 0.947 | 0.811 | 0.807 | 0.42 | 0.341 | 0.750 | incalculable | 0.600 | 0.364 | 0.828 | incalculable | 0.764 | 0.455 | incalculable | 0.921 |
| ODDPub | Sensibility (CI 95%) |  |  | 0.26 (0.13 - 0.44) | 0.19 (0.04 - 0.46) |  | 0.38 (0.09 - 0.76) |  |  |  |  |  |  |  |  |  |
|  | Specificity (CI 95%) |  |  | incalculable | 0.94 (0.73 - 1) |  | 0.88 (0.70 - 0.98) |  |  |  |  |  |  |  |  |  |
|  | F1-score |  |  | 0.419 | 0.30 |  | 0.429 |  |  |  |  |  |  |  |  |  |
| SciScore | Sensibility (CI 95%) |  | 0.05 (0.00 - 0.26) | 0.42 (0.25 - 0.61) |  |  | 0.29 (0.04 - 0.71) |  |  |  |  |  |  |  |  |  |
|  | Specificity (CI 95%) |  | 1.00 (0.74 - 1.00) | incalculable |  |  | 0.96 (0.79 - 1.00) |  |  |  |  |  |  |  |  |  |
|  | F1-score |  | 0.100 | 0.591 |  |  | 0.400 |  |  |  |  |  |  |  |  |  |
| TNRscreener | Sensibility (CI 95%) |  | 0.43 (0.22 - 0.66) |  |  |  |  |  |  |  |  |  |  |  |  |  |
|  | Specificity (CI 95%) |  | 0.77 (0.46 - 0.95) |  |  |  |  |  |  |  |  |  |  |  |  |  |
|  | F1-score |  | 0.545 |  |  |  |  |  |  |  |  |  |  |  |  |  |
| rTransparent | Sensibility (CI 95%) |  | 0.90 (0.70 - 0.99) | 0.32 (0.17 - 0.51) | 0.19 (0.04 - 0.46) |  | 0.50 (0.16 - 0.84) |  |  |  |  |  |  | 1.00 (0.90 - 1.00) |  | 1.00 (0.90 - 1.00) |
|  | Specificity (CI 95%) |  | 1.00 (0.75 - 1.00) | incalculable | 0.83 (0.59 - 0.96) |  | 0.96 (0.80 - 1.00) |  |  |  |  |  |  | incalculable |  | incalculable |
|  | F1-score |  | 0.950 | 0.489 | 0.27 |  | 0.615 |  |  |  |  |  |  | 1.000 |  | 1.000 |
| Unpaywall | Sensibility (CI 95%) |  |  |  |  | 1 (0.90-1) |  |  |  |  |  |  |  |  |  |  |
|  | Specificity (CI 95%) |  |  |  |  | incalculable |  |  |  |  |  |  |  |  |  |  |
|  | F1-score |  |  |  |  | 1 |  |  |  |  |  |  |  |  |  |  |

**Web Appendix 25: Justification for each open science practice of the chosen tool:**

*When a commercial tool exists, the fact that the tool is open access was also a reason, if the performances are comparable.*

| Practice | Tool selected | Reason |
| --- | --- | --- |
| Registration | rTransparent | One of the best performances (2^nd^ one), easy to use, open access |
| Data sharing | Llama 3.3 70B | Best performance, open access |
| Open access publishing | Unpaywall | Best performance |
| Code sharing | Llama 3.3 70B | Best performance, open access |
| Publication 1year (for RCT) | Llama 3.3 70B | Only option |
| Protocol | Llama 3.3 70B | Only option |
| SAP | Llama 3.3 70B | Only option |
| Reporting Guideline | Llama 3.3 70B | Only option |
| Preprint | Llama 3.3 70B | Best performance, open access |
| Author contribution | ContriBOT | Best performance |
| COI | rTransparent | Best performance |
| ORCID | ContriBOT | Best performance |
| Funding statement | rTransparent | Best performance |

**Web Appendix 26: Automated tool performance in XML and PDF files.**

rTransparent:

| Type of files | Metric | Registration | COI | Funding statement |
| --- | --- | --- | --- | --- |
| XML | Sensibility (CI 95%) | 0.01 (0.00 - 0.05) | 1.00 (0.98 -1.00) | 0.98 (0.94 - 0.99) |
|  | Specificity (CI 95%) | 1.00 (0.95 - 1.00) | incalculable | 0.75 (0.19 - 0.99) |
|  | F1-score | 0.018 | 1.00 | 0.986 |
| PDF | Sensibility (CI 95%) | 0.90 (0.83 – 0.95) | 1.00 (0.98 - 1.00) | 0.98 (0.94 - 0.99) |
|  | Specificity (CI 95%) | 0.90 (0.80 – 0.96) | incalculable | 0.50 (0.07 - 0.93) |
|  | F1-score | 0.92 | 1.00 | 0.98 |

ContriBOT:

| Type of files | Metric | Contribution of the author | ORCID |
| --- | --- | --- | --- |
| XML | Sensibility (CI 95%) | 0.00 (0.00 – 0.02) | 0 .00 (0.00 – 0.03) |
|  | Specificity (CI 95%) | 1.00 (0.03- 1.00) | 1.00 (0.94 – 1.00) |
|  | F1-score | incalculable | incalculable |
| PDF | Sensibility (CI 95%) | 0.99 (0.97 – 1.00) | 098 (0.93 – 1.00) |
|  | Specificity (CI 95%) | 1.00 (0.3 – 1.00) | 1.00 (0.95 – 1.00) |
|  | F1-score | 1.00 | 0.99 |

Llama 3.3 70B:

| Tool | Metric | Randomisation | Data Sharing | Open access publishing | Code sharing | Publication 1year (for RCT) | Protocol | SAP | Reporting Guideline | Preprint |
| --- | --- | --- | --- | --- | --- | --- | --- | --- | --- | --- |
| XML | Sensibility (CI 95%) | 1.00 (0.93 - 1.00) | 0.83 (0.76 - 0.89) | 0.99 (0.97 - 1.00) | 0.78 (0.62 - 0.89) | 0.15 (0.02 - 0.45) | 0.80 (0.71 - 0.88) | 0.94 (0.73 - 1.00) | 0.96 (0.90 - 0.99) | incalculable |
|  | Specificity (CI 95%) | 0.93 (0.87 - 0.97) | 0.54 (0.34 - 0.72) | incalculable | 0.77 (0.70 - 0.84) | 0.98 (0.89 - 1.00) | 0.46 (0.35 - 0.57) | 0.57 (0.49 - 0.64) | 0.74 (0.62 - 0.84) | 0.93 (0.88 - 0.96) |
|  | F1-score | 0.92 | 0.87 | 1.00 | 0.60 | 0.25 | 0.71 | 0.32 | 0.90 | incalculable |
| PDF | Sensibility (CI 95%) | 1.00 (0.93 - 1.00) | 0.81 (0.73 - 0.86) | 0.35 (0.28 - 0.42) | 0.72 (0.56 - 0.85) | 0.08 (0.00 - 0.36) | 0.66 (0.56 - 0.75) | 0.89 (0.65 - 0.99) | 0.65 (0.56 - 0.74) | incalculable |
|  | Specificity (CI 95%) | 0.95 (0.89 - 0.98) | 0.61 (0.41 - 0.78) | incalculable | 0.78 (0.70 - 0.85) | 1.00 (0.93 - 1.00) | 0.46 (0.35 - 0.57) | 0.43 (0.35 - 0.51) | 0.84 (0.73 - 0.92) | 0.98 (0.94 - 0.99) |
|  | F1-score | 0.94 | 0.86 | 0.51 | 0.58 | 0.14 | 0.62 | 0.25 | 0.75 | incalculable |
